# Type I interferon responses contribute to immune protection against mycobacterial infection

**DOI:** 10.1101/2024.06.26.24309490

**Authors:** Andrea Szydlo-Shein, Blanca Sanz-Magallón Duque de Estrada, Joshua Rosenheim, Carolin T. Turner, Aneesh Chandran, Evdokia Tsaliki, Marc C. I. Lipman, Heinke Kunst, Gabriele Pollara, Philip M. Elks, Jean-Pierre Levraud, Elspeth M. Payne, Mahdad Noursadeghi, Gillian S. Tomlinson

**Author notes:** **Corresponding author** Gillian S. Tomlinson.

## Abstract

Type I interferon responses have been considered detrimental to host protection in tuberculosis (TB). We provide novel data to challenge this paradigm, derived from transcriptional profiling of human in vivo immune responses to discover associations with radiographic disease severity in pulmonary TB, combined with mechanistic studies to test causality for observed associations using a zebrafish larval mycobacterial infection model. Type I interferon activity in tissue samples from the site of a standardised mycobacterial challenge, the tuberculin skin test, was associated with less severe human TB disease. Abrogation of type I interferon signalling, by CRISPR-mediated mutagenesis of *stat2*, led to increased burden and dissemination of *Mycobacterium marinum* infection in zebrafish larvae. The mechanism for increased severity of mycobacterial infection in zebrafish involves reduced recruitment of myeloid cells required to restrict bacterial growth. Our data support a clear host protective role for type I interferon responses in mycobacterial infection, with potential applications for risk-stratification of adverse outcomes and development of a host-directed therapy to mitigate against severe disease.

## Introduction

*Mycobacterium tuberculosis* (Mtb) infection results in diverse clinical manifestations, ranging from asymptomatic infection to active tuberculosis (TB), which comprises an enormous spectrum of disease severity ^1,2^. Although we know that immune responses contribute to the risk of disease, current understanding of the role of immune response variation in disease severity is limited. Identification of immune correlates of disease severity may offer new insights into the immune pathways that contribute to protection and pathogenesis. This is necessary to enable novel vaccine design and evaluation, stratify disease risk in people who become infected and inform development of novel host-directed therapies to shorten treatment regimens, mitigate against antimicrobial resistance, and against the chronic sequelae of tissue injury in TB ^3^.

To date, protective immunity has focussed on tumour necrosis factor (TNF) and type II interferon mediated immune signalling pathways ^4^. Type I interferon immune responses are also pervasive in TB. Unlike the protective role of type I interferons in viral infections, the prevailing view has been that this pathway may contribute to TB pathogenesis ^5^. However, causal inferences have been limited to mouse models which do not represent a natural host-pathogen interaction ^6^. We sought to re-evaluate the differential role of immune signalling pathways in protection or pathogenesis in TB, by combining observational human data with genetic perturbation of immune signalling pathways in zebrafish infected with its natural pathogen *Mycobacterium marinum* (Mm).

Observational immunology in human TB is limited by reverse causality. This is particularly true for positive correlations between immune responses and disease severity. Therefore, we specifically sought to identify negative correlations with disease severity, that are less susceptible to this limitation and suggestive of protective immunity. Transcriptional profiling can be used to make unbiased system level assessment of immune signalling pathways. In patients with TB, we used transcriptional profiling to investigate steady state perturbation of the immune system in peripheral blood ^7^, and to measure inducible immune responses at the tissue site of a standardised Mtb challenge modelled by the tuberculin skin test (TST) ^8,9^. Our innovative strategy revealed an inverse relationship between type I interferon activity in the TST transcriptome and radiographic severity of human TB, challenging the paradigm of a host-pathogenic role for type I interferons. Here, we confirmed a role for type I interferon responses in host protection against mycobacteria by showing that mutagenesis of *stat2* to abrogate downstream type I interferon signalling, led to increased severity of *M. marinum* infection in zebrafish larvae, due to reduced recruitment of macrophages and neutrophils required to contain bacterial growth.

## Results

### Immune pathways negatively correlated with TB disease severity are not evident in the steady state peripheral blood transcriptome

Our analysis of human immune responses in TB leveraged a previously reported cohort of patients with pulmonary TB ^10^ which we have extended to a total sample size of 51 people with microbiologically confirmed disease, within one month of commencing anti-tuberculous chemotherapy (Table 1, Figure S1). We evaluated disease severity using a published radiographic scoring system which incorporates extent of disease and the presence of cavitation on chest x-ray ^11^. We performed genome-wide transcriptional profiling of peripheral blood, and punch biopsies from the site of a 48 hour TST. Transcriptional changes associated with the immune response to TB were identified by comparison with reference healthy control blood samples and punch biopsies from the site of control saline injections. We used outlier profile analysis ^12^ to capture inter-individual variation in immune perturbation associated with active TB. This overcomes the limitation of conventional differential gene expression analysis that may exclude changes that occur in only a subset of cases. In outlier profile analysis, we identified transcripts in the peripheral blood of each individual that were significantly enriched (more abundant) compared to control blood samples from healthy volunteers. The integrated list of outlier genes from the entire cohort defined an active TB blood transcriptome comprised of 2620 genes, enriched for pathways involved in innate and adaptive immune responses and cell cycle regulation (Figure S2).

**Table 1.** Clinical and demographic data summary. Demographic characteristics and magnitude of clinical induration of the skin at the site of TST or saline injection are summarised for all study subjects. Duration of anti-tuberculous drug treatment at the time of sampling and radiographic disease severity data are provided for individuals with pulmonary TB. *Peripheral blood samples were also taken from 50 of these individuals with pulmonary TB. Saline samples were obtained from 17 individuals with pulmonary TB and 32 healthy participants who were people with cured or latent TB, BCG vaccine recipients and healthy volunteers, described previously ^10^. Control blood samples from healthy individuals were described previously ^13^. **Applies only to participants with active TB receiving saline injections. CXR = chest x-ray.

|  |  | TST* | Saline controls | Blood controls |
| --- | --- | --- | --- | --- |
| Number |  | 51 | 49 | 58 |
| Age | (median and range) | 30 (19-77) | 28 (18-75) | 36 (21-69) |
| Sex | Male | 31 (61%) | 19 (39%) | 16 (28%) |
|  | Female | 20 (39%) | 30 (61%) | 42 (72%) |
| Ethnicity | White | 21 (41%) | 20 (41%) | 44 (76%) |
|  | Asian | 15 (29%) | 8 (16%) | 9 (15%) |
|  | Black | 12 (24%) | 7 (14%) | 1 (2%) |
|  | Mixed | 2 (4%) | 1 (2%) | 4 (7%) |
|  | South American | 1 (2%) | 11 (22%) | 0 |
|  | Not recorded | 0 | 2 (4%) | 0 |
| Intradermal injection site induration (mm) | (median and range) | 20 (0-70) | 0 | NA |
| Days TB treatment before TST +/- saline | (median and range) | 19 (0-30) | 15 (0-28)** | NA |
| CXR severity score | (median and range) | 30 (0-140) | 15 (0-110)** | NA |

Next, we sought to identify immunological pathways correlated with disease severity. We identified individual genes in the blood transcriptome that showed statistically significant correlations with the radiographic severity score (Figure 1A). Pathway analysis revealed no enrichment at pathway level among genes negatively correlated with TB severity. There was modest enrichment for innate and adaptive immune response pathways among genes positively correlated with radiographic disease score (Figure 1B). Next, we subjected these genes to Ingenuity Pathway Analysis (IPA) upstream regulator analysis to collate groups of co-regulated genes (Figure 1A and Figure S3A). These were annotated by their upstream regulators at the level of cytokines, transmembrane receptors, kinases and transcriptional regulators, representing the key components of signalling pathways which mediate immune responses. To provide increased confidence that these modules represented co-regulated genes in each molecular pathway, we retained only those that had significantly greater co-correlated expression than modules of genes randomly selected from our blood transcriptomes ^13^. Only six gene modules were identified among genes negatively correlated with disease severity, but these did not represent obvious coordinated biology (Figure S3B). Among genes positively correlated with TB severity 94 gene modules were identified, reflecting pro-inflammatory cytokine activity, type II interferon and T cell activation and innate immune signalling pathways (Figure S3C and Figure 1C).

**Figure 1.**
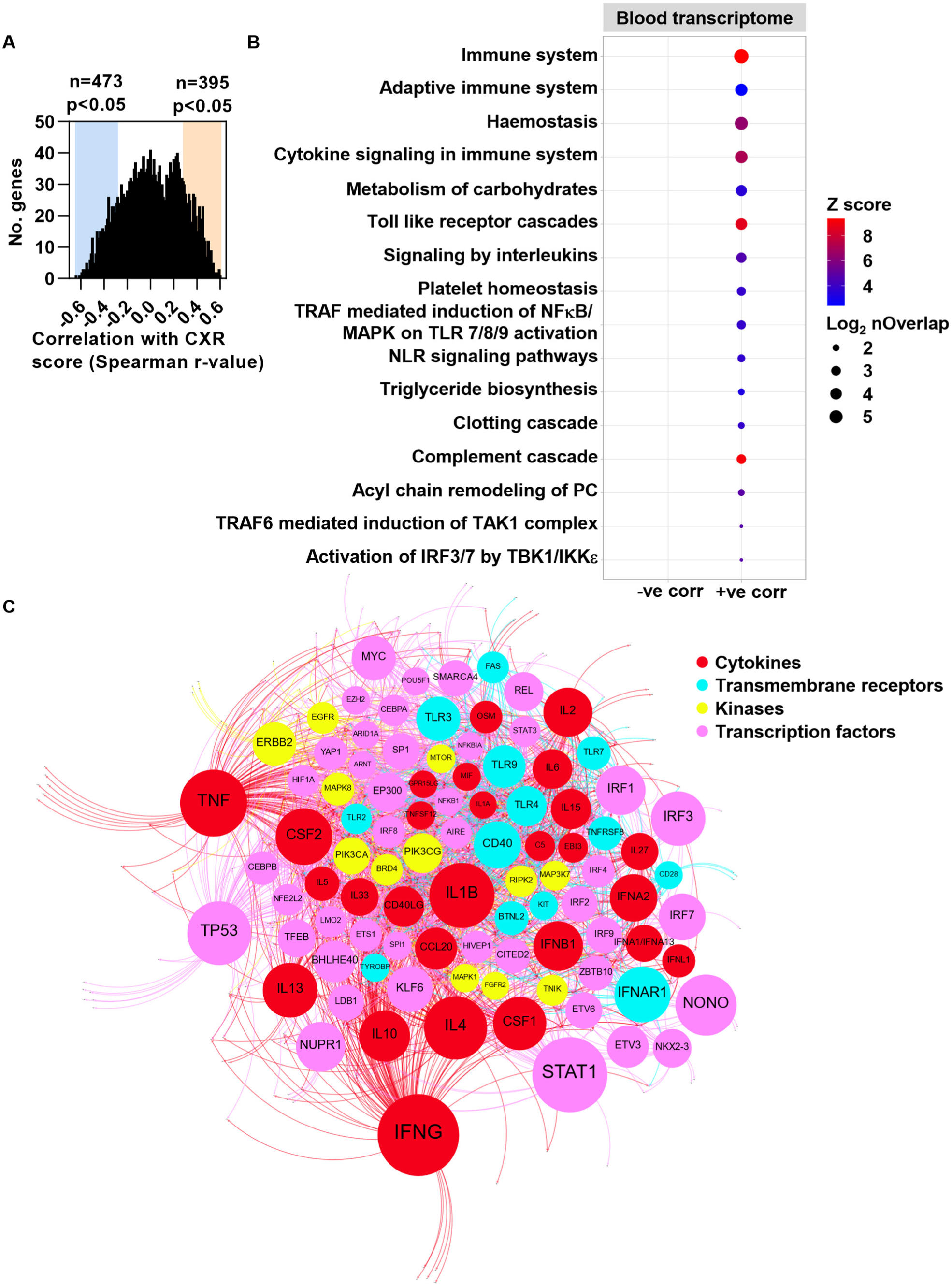
The steady state human peripheral blood transcriptome reflects immune responses positively correlated with TB severity. **(A)** Frequency distribution of the correlation coefficients for the relationship between the abundance of each of the 2620 transcripts which define the peripheral blood transcriptome derived from 50 individuals with pulmonary tuberculosis (TB) and radiographic disease severity. Coloured boxes highlight statistically significant correlations (p<0.05). **(B)** Reactome pathway enrichment among blood transcriptome genes whose expression is statistically significantly correlated with radiographic TB disease severity; corr = correlated. **(C)** Network diagram of statistically significant (FDR<0.05) upstream regulators (labelled nodes) coloured by their molecular function, of target gene modules whose expression is positively correlated with radiological TB severity. Size of the nodes for upstream regulators is proportional to -Log10 adjusted p value. Nodes were clustered using the Force Atlas 2 algorithm in Gephi (version 0.9.2).

### Outlier analysis of the TST transcriptome defines inducible human in vivo immune responses in active tuberculosis

We repeated the same analysis of the dynamic response to TST challenge in the same patient cohort. In this analysis, we identified transcripts in the TST of each individual that were significantly enriched compared to gene expression in control data from skin biopsies at the site of saline injections. The integrated list of outlier genes statistically enriched in TST samples from all 51 individuals compared to control saline samples defined an active TB TST transcriptome comprised of 3222 transcripts. This analysis revealed that although some aspects of the TST response are consistent in all people, there is also considerable variability between individuals (Figure S4A). The 3222 transcripts identified by this approach, were enriched for canonical pathways involved in cell mediated responses, consistent with our previous description of the TST gene signature identified by conventional differential gene expression analysis ^9,10^ (Figure S4B).

### Reduced type I interferon activity in the TST transcriptome is associated with increased severity of human TB disease

In contrast to the peripheral blood analysis, pathway analysis of individual genes in the TST transcriptome that showed statistically significant correlations with the radiographic severity score identified enrichment for cell mediated immune responses among genes negatively correlated with disease severity. Statistically less significant enrichment for immune response pathways was evident among genes positively correlated with TB severity (Figure 2A and Figure S5A). Subsequent IPA upstream regulator analysis identified 111 modules among genes inversely correlated with radiographic TB severity, suggesting they may contribute to host protection against severe disease (Figure 2A-C and Figure S5B). These included transcriptional modules representing pro-inflammatory cytokine activity, interferon gamma and T cell activity, consistent with existing evidence predominantly supporting their protective roles in TB ^4^ (Figure 2B). In addition, we found enrichment of multiple co-regulated gene networks attributed to type I interferon signalling, suggesting that type I interferon responses may also contribute to host protection in mycobacterial infection (Figure 2B, C). Interferon gamma was also predicted to regulate one of only two small putative networks within genes positively correlated with disease severity; the other driven by TCL1A, an AKT kinase activator that promotes cellular survival ^14^ (Figure S5C).

**Figure 2.**
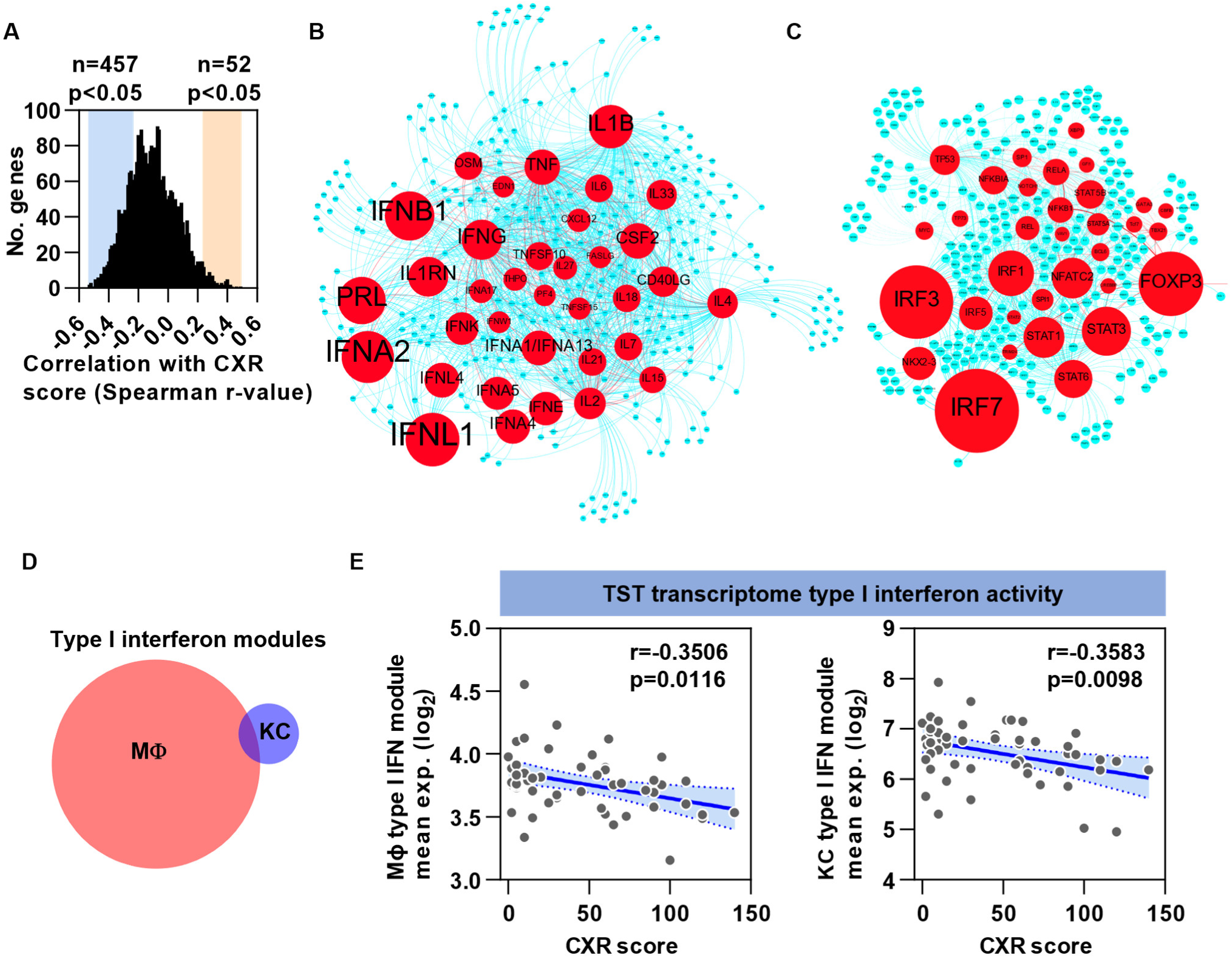
Reduced type I interferon activity in the TST transcriptome is associated with increased severity of human pulmonary TB disease. **(A)** Frequency distribution of the correlation coefficients for the relationship between the abundance of each of the 3222 transcripts within the TST transcriptome derived from 51 individuals with pulmonary TB and radiographic disease severity. Coloured boxes highlight statistically significant correlations (p<0.05). **(B, C)** Network diagrams of statistically significant (FDR<0.05) cytokines **(B)** and transcription factors **(C)** predicted to be upstream regulators (red) of target gene (blue) modules whose expression is negatively correlated with radiological TB severity. Size of the nodes for upstream regulators is proportional to -Log10 adjusted p value. Nodes were clustered using the Force Atlas 2 algorithm in Gephi (version 0.9.2). **(D, E)** The inverse relationship between average expression in each TST sample of two distinct, independently derived gene modules specific for type I interferon activity (Table S1) **(D)** with TB severity **(E)**. r values and p values were derived from two-tailed Spearman rank correlations. CXR = Chest x-ray, MΦ = Macrophage, KC = Keratinocyte.

To validate the immunological responses predicted to be associated with disease severity by upstream regulator analysis, we quantified the expression of two independently derived and largely non-overlapping type I interferon inducible gene modules reflecting the specific biological activity of type I interferons (Table S1) ^9,10^, within the TB TST transcriptome (Figure 2D). This provided independent validation of the inverse relationship between type I interferon responses and radiographic severity of TB (Figure 2E), suggestive of a host-protective role. By contrast, the expression of the entire TST transcriptome and that of three distinct independently derived modules specific for interferon gamma inducible genes ^9^ (Table S1) did not correlate with disease severity (Figure S6A-E). There was also no relationship between peripheral blood type I IFN activity and radiological severity score (Figure S6F, G).

### *stat2* CRISPant zebrafish larvae exhibit increased susceptibility to *M. marinum* infection

The finding of numerous modules pertaining to type I interferon signalling among TST transcriptome genes negatively correlated with disease severity prompted us to test the role of this pathway in mycobacterial infection experimentally. Since type I interferons are primarily induced following activation of innate immune recognition pathways ^15^, we investigated their role in the zebrafish larval *M. marinum* infection model, which allows exclusive assessment of innate immunity ^16^. The mammalian canonical type I interferon signalling pathway is highly conserved in zebrafish ^17,18^ (Figure 3A). First, we confirmed that intravenous *M. marinum* infection of wild type zebrafish larvae induces a type I interferon response, by showing enrichment for two distinct independently derived transcriptional modules reflecting type I interferon activity ^13,18^ (Table S1) within genome-wide transcriptomic data from whole embryos after four days of mycobacterial growth (Figure 3B, C). To block downstream type I interferon signalling we used CRISPR-mediated mutagenesis of *stat2*, as a critical component of the principal transcription factor complex which induces expression of type I interferon stimulated genes (ISG) ^19–21^, for which there is a single zebrafish orthologue ^17^ (Figure 3A). Injection of three guide RNA/Cas9 ribonucleoprotein (RNP) complexes each targeting a distinct *stat2* exon, into one cell stage wild type zebrafish embryos led to >90% efficient mutagenesis in CRISPant embryos, confirmed by next-generation sequencing (Figure S7A and Figure S8A). We verified loss of functional downstream type I interferon signalling by showing that induction of the classical ISG Mxa ^18,21^ by injection of recombinant Interferon phi 1 protein, measured by expression of a reporter *mxa:mCherry* transgene ^22^, was significantly attenuated in *stat2* CRISPants compared to siblings injected with “scrambled” (negative control) RNPs designed to lack genomic targets (Figure S7B and Figure S8B-C). *stat2* CRISPants developed normally up to five days post-fertilisation (Figure S8B).

**Figure 3.**
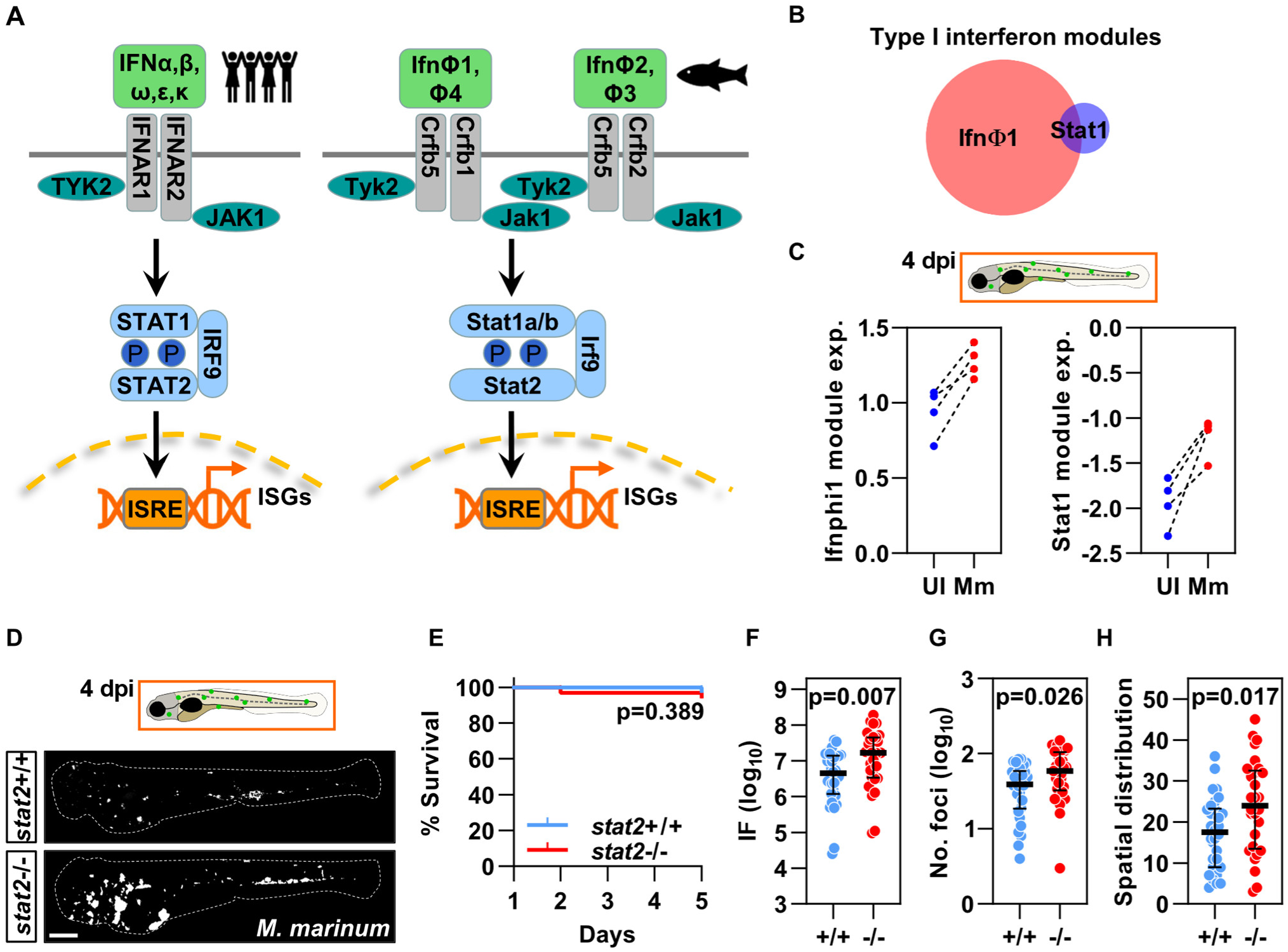
Increased severity of *M. marinum* infection in *stat2* CRISPants. **(A)** Schematic representation of the human canonical type I interferon signalling pathway (left) which is highly conserved in zebrafish (right). **(B, C)** The average expression of two largely non-overlapping **(B)** type I interferon inducible gene modules derived from independent data (Table S1) in genome-wide mRNA sequencing data from whole wild type embryos four days post-intravenous infection with 400 colony forming units (cfu) of *M. marinum* (Mm) compared with uninfected (UI) siblings **(C)**. Data are from four independent experiments. **(D)** *stat2* CRISPant (*stat2*-/-) and control ribonucleoprotein (RNP) injected (*stat2*+/+) zebrafish larvae four days post intravenous infection with 400 cfu *M. marinum*. Scale bar = 250 μm. Dashed white lines indicate the outline of each larva. **(E)** Survival of control and *stat2* disrupted zebrafish larvae 1-4 days post infection with *M. marinum*. p value was derived from a log-rank (Mantel-Cox) test. **(F-H)** Comparison of integrated fluorescence (IF) (a surrogate for bacterial burden) **(F)**, number of bacterial foci **(G)** and spatial distribution of bacteria **(H)**, in *stat2* CRISPants compared to control RNP injected siblings. Each data point represents an individual zebrafish larva, lines and error bars indicate the median and interquartile range. p values were derived from two-tailed Mann-Whitney tests. Data are representative of four experiments. dpi = days post-infection.

There was no difference in survival of *M. marinum* infected *stat2* CRISPants and scrambled RNP injected siblings within the timeframe of our experiments (Figure 3D, E). However, mycobacterial burden, the number of bacterial foci and the spatial dissemination of bacteria, determined by quantitation of fluorescent foci of *M. marinum*, were all significantly higher in intravenously infected *stat2* CRISPants compared to control RNP injected siblings, indicative of more severe disease (Figure 3D, F-H). These findings are consistent with a host protective role for type I interferons in mycobacterial infection, as suggested by our observation of an inverse relationship between type I interferon activity and disease severity in human TB (Figure 2).

### Steady state neutrophil numbers are reduced in *stat2* CRISPant zebrafish larvae

Macrophages are critically important for control of *M. marinum* infection in zebrafish larvae ^23–25^ and neutrophils may also contribute to mycobacterial killing ^26,27^. Therefore, to investigate the mechanisms by which *stat2* deficiency leads to worse mycobacterial infection we used transgenic zebrafish lines with fluorescent immune cell lineages, *Tg(mpeg1:mCherry)* ^28^ and *Tg(mpx:eGFP)* ^29^, to generate *stat2* CRISPants that have fluorescent macrophages or neutrophils, respectively. First, we asked whether *stat2* disruption impacted steady state macrophage or neutrophil numbers. We demonstrated by quantitative fluorescence microscopy, using integrated fluorescence as a surrogate for cell number, that *stat2* deficiency did not affect baseline macrophage abundance (Figure 4A, B). However, it was associated with reduced neutrophil abundance (Figure 4C, D), a phenotype also reported in human STAT2 deficiency ^30^.

**Figure 4.**
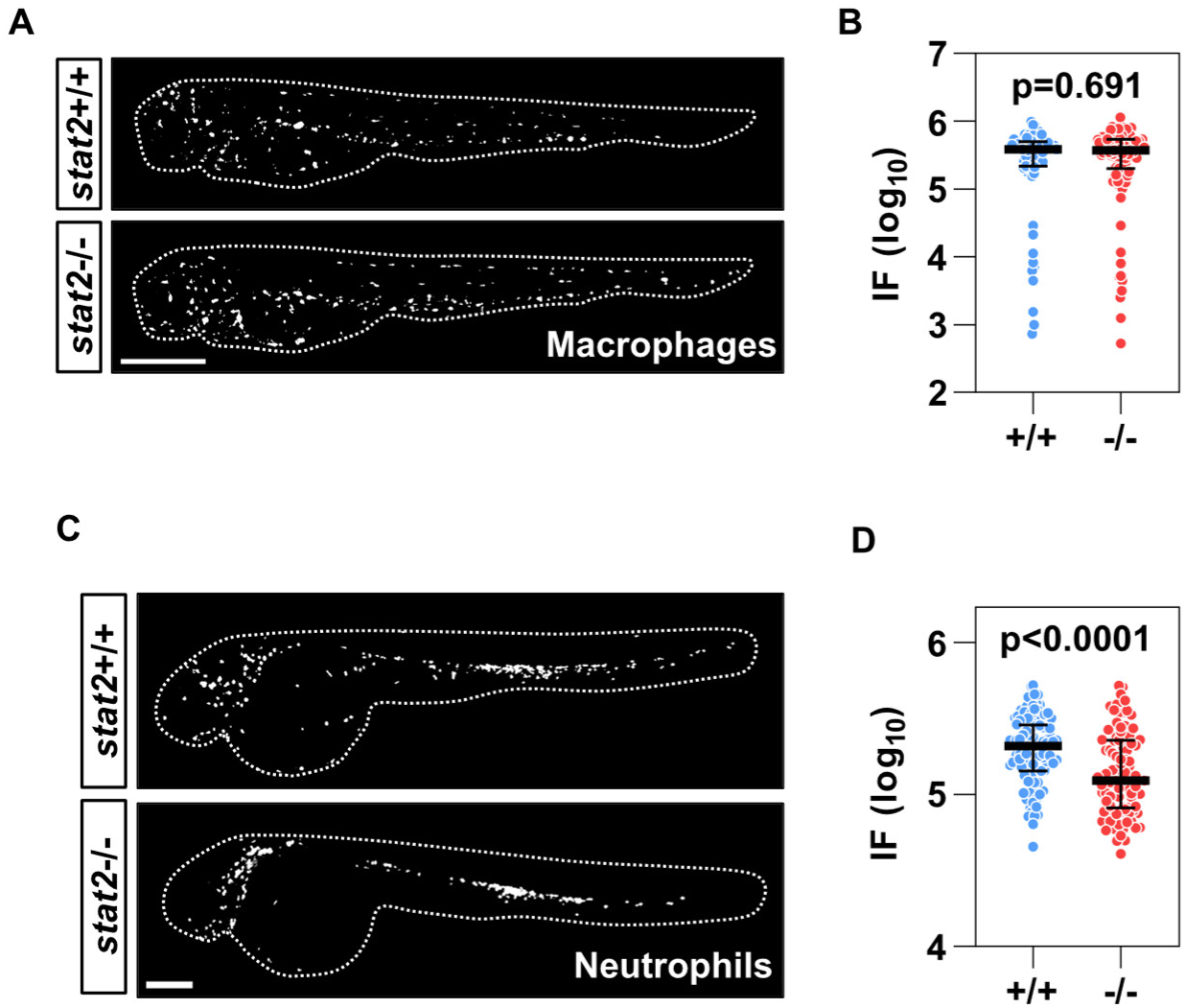
Reduced steady state neutrophil numbers in *stat2* CRISPants. **(A)** Representative images of three day post-fertilisation (dpf) control (upper panel) and *stat2* CRISPant (lower panel) *Tg(mpeg1:mCherry)* zebrafish larvae delineated by the dashed white lines. Scale bar = 250 μm. **(B)** Integrated fluorescence (IF) as a surrogate for steady state macrophage numbers in three dpf *stat2* CRISPants compared to siblings injected with scrambled ribonucleoproteins (RNPs). **(C)** Representative images of two dpf control (upper panel) and *stat2* CRISPant (lower panel) *Tg(mpx:eGFP)* zebrafish larvae delineated by the dashed white lines. Scale bar = 250 μm. **(D)** Quantitation of baseline neutrophil numbers (represented by IF) in two dpf *stat2* CRISPants and negative control RNP injected siblings. Each data point represents an individual zebrafish larva, lines and error bars indicate the median and interquartile range. p values were derived from two-tailed Mann-Whitney tests. Data are from three independent experiments.

### Recruitment of macrophages but not neutrophils to the site of sterile injury is *stat2* dependent

Type I interferons have previously been shown to influence cellular migration ^15,31^. Therefore, we tested macrophage and neutrophil recruitment in *stat2* CRISPants using a tailfin transection model of sterile injury, most commonly used to evaluate neutrophil recruitment ^29^ (Figure S7C). The timing of neutrophil homing to the site of sterile tail fin transection was normal in *stat2* CRISPants, with maximal accumulation at six hours and dissipation of recruited cells becoming evident by 24 hours, similar to that of control RNP injected embryos (Figure 5A, B and Figure S9) and in keeping with previous studies ^29,32^. Despite baseline neutropenia, the number of neutrophils at the wound site at the point of maximal recruitment in *stat2* CRISPants exceeded that of control RNP injected embryos. But by 24 hours the number of neutrophils at the injury site was significantly reduced in *stat2* CRISPants, likely reflecting the attenuated increase in total neutrophil numbers in response to the acute inflammatory challenge (Figure 5C, D).

**Figure 5.**
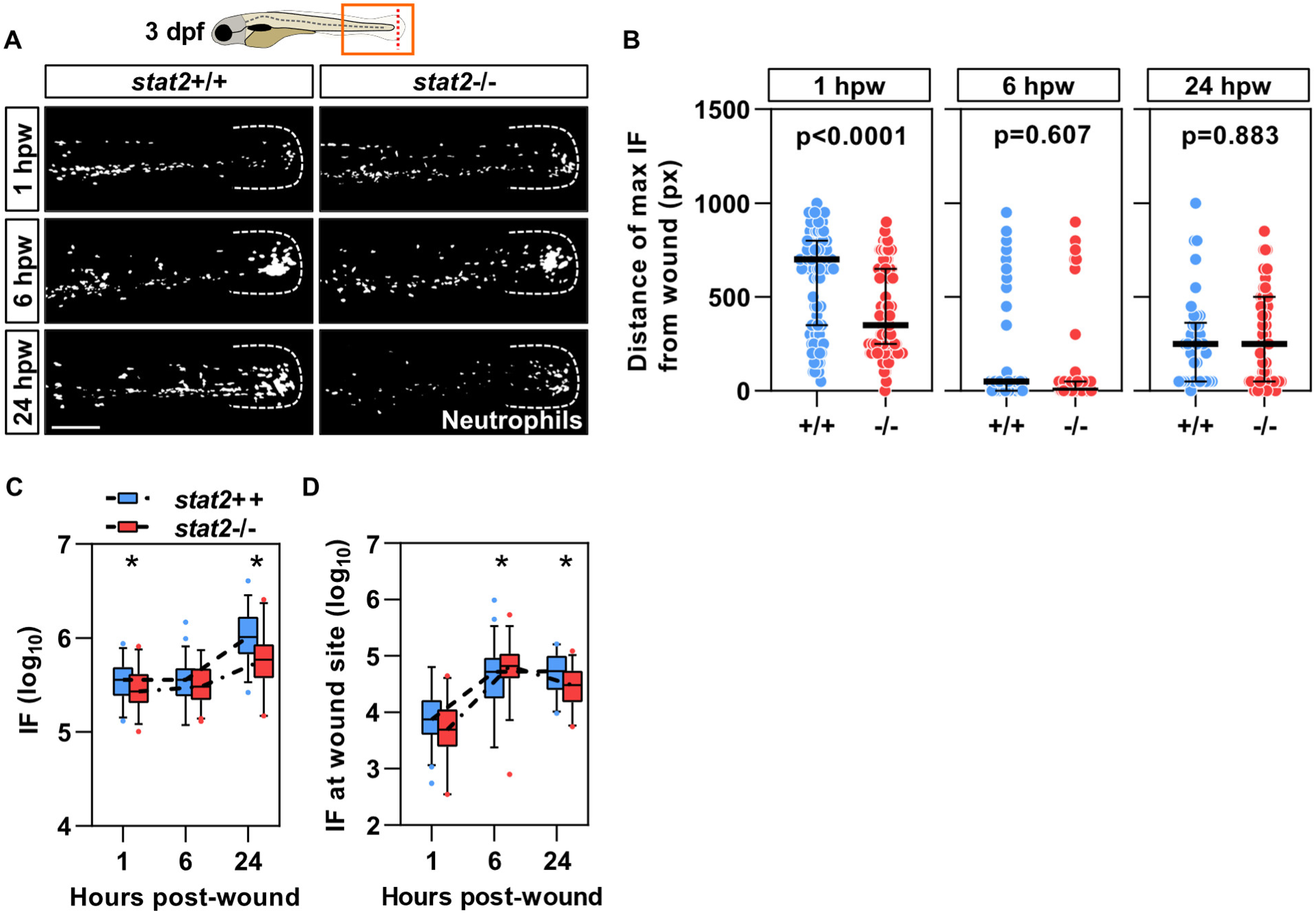
Neutrophil recruitment to the site of sterile injury is not *stat2* dependent. **(A)** Representative images of eGFP-expressing neutrophils recruited to the site of injury 1, 6 and 24 hours following tailfin transection of three day post-fertilisation (dpf) *stat2* CRISPant *Tg(mpx:eGFP)* zebrafish larvae and scrambled ribonucleoprotein (RNP) injected controls. Scale bar represents 100 μm. The white dashes indicate the outline of the tailfin. **(B)** Distance of neutrophils from the injury site at 1, 6 and 24 hours post-wound (hpw), derived by identifying the Sholl circle in which the maximum integrated fluorescence (IF), a surrogate for highest cell numbers, was detected in *stat2* CRISPants and scrambled RNP injected siblings. **(C-D)** IF, a surrogate for total neutrophil numbers **(C)**, and IF at the wound site, representing neutrophil numbers localized to the tailfin transection **(D)** are shown at 1, 6 and 24 hours following injury. On scatter plots data points represent individual zebrafish larvae and lines and error bars the median and interquartile range. On box and whisker plots horizontal lines represent median values, box limits indicate the interquartile range and whiskers extend between the 5th and 95th percentiles. p values were derived from two-tailed Mann-Whitney tests. * = p<0.05. Data are from three independent experiments. px = Pixels.

Macrophage recruitment to the site of a sterile tail wound has previously been reported to be slower than that of neutrophils ^32,33^. Very few macrophages were present at the tailfin transection site by one hour in either *stat2* disrupted or control transgenic larvae, but recruitment progressively increased until 24 hours in both groups (Figure 6). Fewer macrophages were found in the vicinity of the wound site at all time points post-injury in *stat2* CRISPant transgenic embryos compared to control RNP injected siblings, the majority being located outwith the tailfin (Figure 6). Taken together these data suggest that *stat2* is necessary for macrophage recruitment but non-essential for neutrophil recruitment to a sterile injury site. In addition, baseline neutropenia did not impact the number of cells localised to the wound at peak neutrophil recruitment.

**Figure 6.**
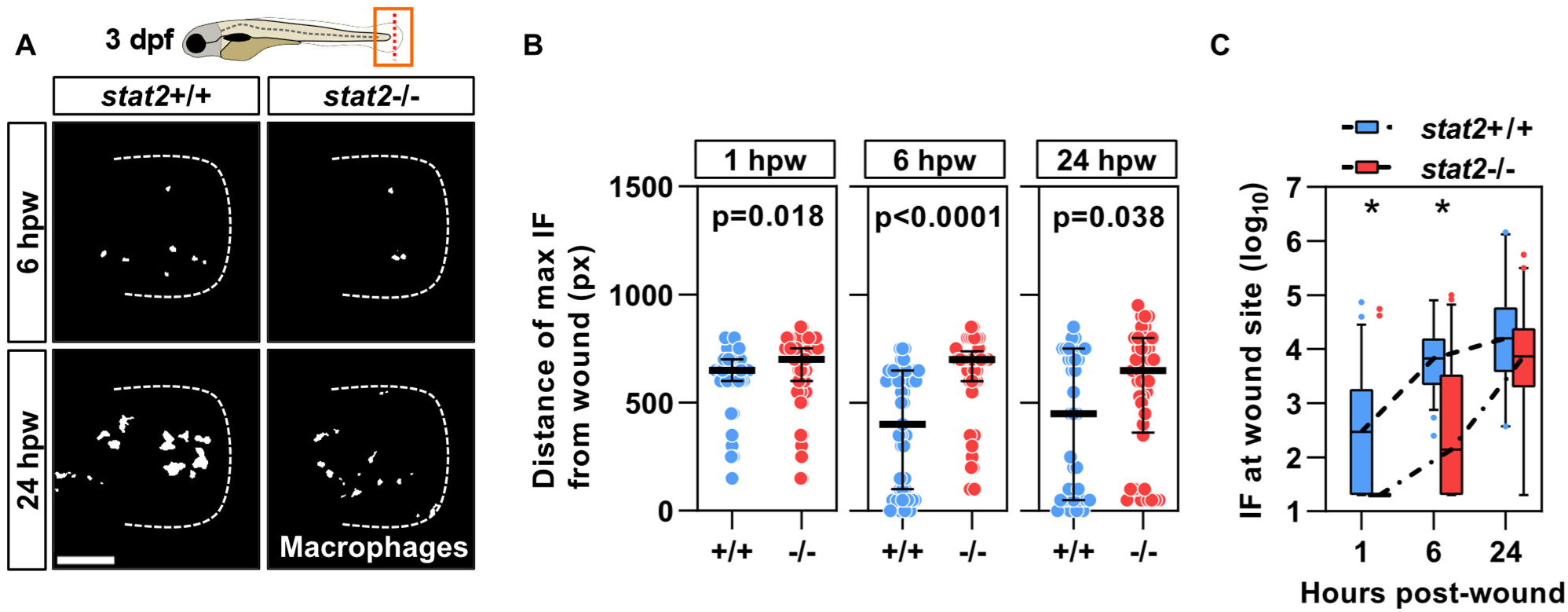
Reduced macrophage recruitment to the site of sterile injury in *stat2* CRISPants. **(A)** Representative images of mCherry-expressing macrophages recruited to the site of tailfin transection by 6 and 24 hours, in three day post-fertilisation (dpf) *stat2* CRISPant *Tg(mpeg1:mCherry)* zebrafish embryos and controls. Scale bar represents 100 μm. The dashed white lines indicate the edge of the tailfin. **(B)** Comparison of the distance of macrophages from the site of injury at the indicated time points for *stat2* CRISPants and control RNP injected siblings. **(C)** Integrated fluorescence (IF) a surrogate for macrophage numbers at the site of tailfin transection is shown at 1, 6 and 24 hours post-wound (hpw). On scatter plots data points represent individual zebrafish larvae and lines and error bars the median and interquartile range. On box and whisker plots horizontal lines represent median values, box limits indicate the interquartile range and whiskers extend between the 5th and 95th percentiles. p values were derived from two-tailed Mann-Whitney tests. * = p<0.05. Data are from three independent experiments. px = Pixels.

### Recruitment of myeloid cells to the site of *M. marinum* infection is *stat2* dependent

Next we focused on macrophage recruitment to the site of mycobacterial infection because this has been reported to be crucial for control of *M. marinum* growth in zebrafish larvae ^24^. We found a significant reduction in the number of macrophages recruited to the site of localised hindbrain *M. marinum* infection in *stat2* deficient transgenic zebrafish larvae compared to their counterparts injected with scrambled RNPs (Figure S7D and Figure 7A, B). We then evaluated whether *stat2* disruption affected recruitment of neutrophils to the site of mycobacterial infection. We found that in contrast to their normal peak recruitment to a sterile wound, the number of neutrophils attracted to the site of mycobacterial disease was significantly diminished in *stat2* CRISPants (Figure 7C, D). Collectively, these results demonstrate that myeloid cell recruitment to the site of *M. marinum* infection is *stat2* dependent.

**Figure 7.**
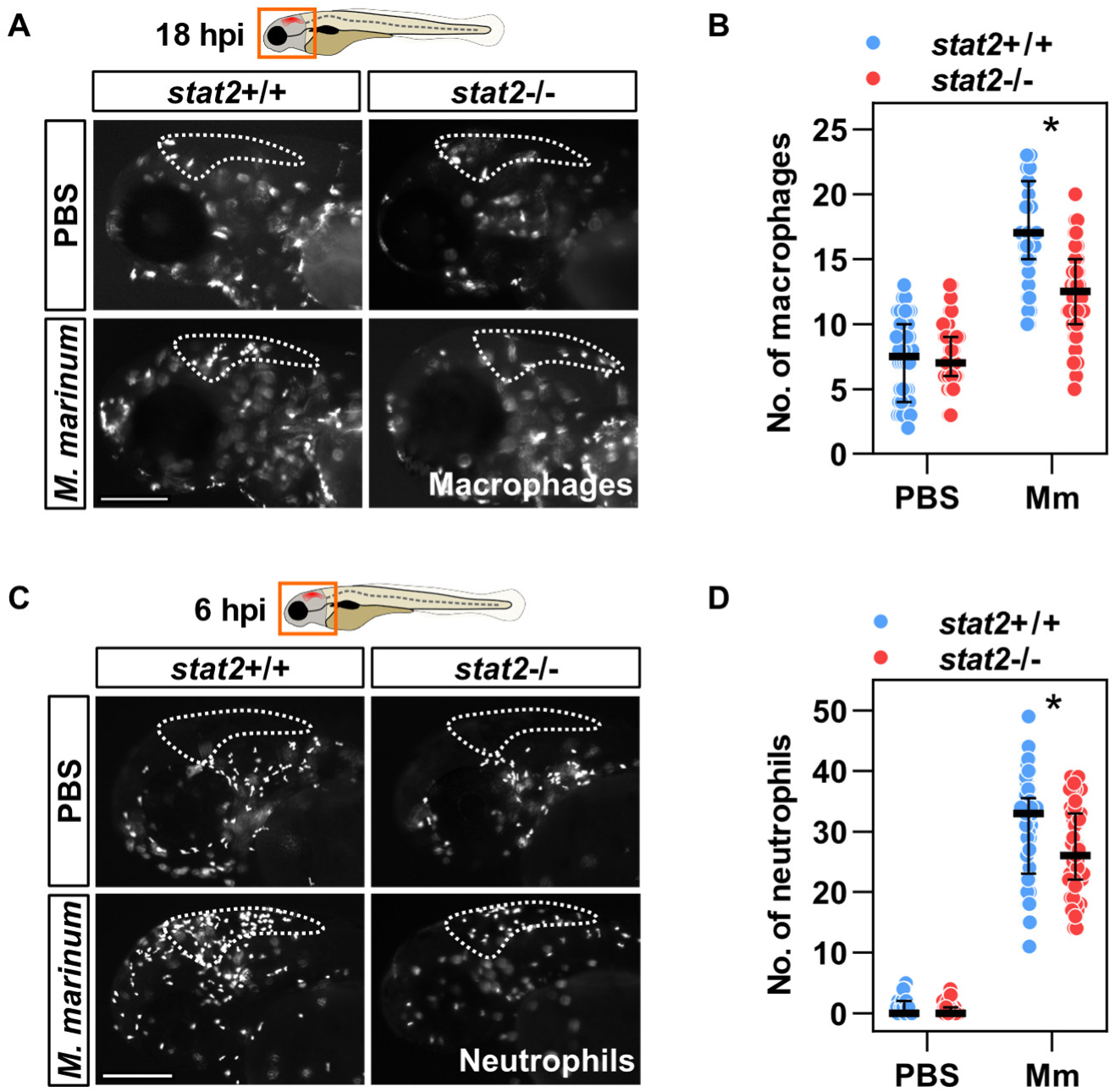
Reduced macrophage and neutrophil recruitment to *M. marinum* in *stat2* CRISPants. **(A)** Representative images of mCherry-expressing macrophages recruited to the hindbrain (dashed white outline) 18 hours post-injection (hpi) of PBS or 200 colony forming units (cfu) *M. marinum* in two day post-fertilisation (dpf) *stat2* CRISPant *Tg(mpeg1:mCherry)* zebrafish larvae and scrambled ribonucleoprotein (RNP) injected controls. **(B)** Number of macrophages recruited to the hindbrain in response to PBS or *M. marinum* in *stat2* CRISPants and negative control RNP injected siblings. **(C)** Representative images of eGFP-expressing neutrophils recruited to the hindbrain (dashed white outline) 6 hours after PBS or *M. marinum* injection (200 cfu) in two dpf *stat2* CRISPant and control *Tg(mpx:eGFP)* zebrafish larvae. **(D)** Quantitation of neutrophils recruited to the site of *M. marinum* infection in *stat2* CRISPants and control siblings. Scale bars represent 100 μm. Data points represent individual zebrafish larvae and lines and error bars the median and interquartile range. p values were derived from two-tailed Mann-Whitney tests. * = p<0.05. Data are from three independent experiments.

In total, the recruitment studies suggest that *stat2* disruption leads to a broad defect in macrophage recruitment to both infectious and non-infectious inflammatory foci. By contrast, *stat2* is non-essential for neutrophil migration to sterile injury. The latter finding is consistent with defective migration, rather than neutropenia, being the principal mechanism for reduced neutrophil numbers at the site of *M. marinum* infection.

## Discussion

We have taken an innovative approach, combining molecular and systems level evaluation of human immune responses to a standardised experimental challenge, to identify biological pathways associated with TB disease severity. We experimentally validated host factors responsible for differences in phenotype using a relevant, tractable animal model which represents a natural host-pathogen relationship ^16^. Molecular interrogation of human in vivo responses to Mtb revealed that reduced type I interferon activity within the TST transcriptome was associated with worse pulmonary disease. Deficient type I interferon signalling, achieved by genetic disruption of *stat2*, led to more severe mycobacterial infection in zebrafish larvae, via a mechanism involving impaired recruitment of macrophages and neutrophils to the site of disease and reduced neutrophil numbers at baseline. Taken together our results demonstrate an important role for type I interferon responses in contributing to innate immune protection against mycobacterial disease.

Previous studies report peripheral blood interferon-inducible gene expression that diminishes with successful treatment in human active TB ^7,34,35^. These transcriptional signatures comprise both type I and type II interferon inducible genes but have mainly been interpreted as type I interferon responses, fostering the view that type I interferons are detrimental to the host in human TB. In the present study we found no relationship between disease severity and peripheral blood expression of gene modules specific for type I interferon activity. Mouse data predominantly suggest that type I interferon responses contribute to increased susceptibility to mycobacterial infection, by mechanisms including antagonism of IL1 mediated protection ^36–38^ and reduced responsiveness of macrophages to interferon gamma ^39^. However, this phenotype has not been universally observed, likely due to differences in host and pathogen genetics ^5^. Moreover, impaired type I interferon responses are generally associated with relatively minor differences in bacterial burden or survival ^5^. Of note, Mtb is not a natural mouse pathogen, and this approach may not model host-pathogen interactions in human TB faithfully ^6^.

In support of a protective role for type I interferons, adjunctive interferon alpha therapy in addition to conventional anti-tuberculous antibiotic treatment led to symptomatic and radiological improvement, with a reduction in bacterial burden in respiratory tract samples, in small-scale human studies ^40–42^. In keeping with this, an early peripheral blood type I interferon-inducible gene signature predicts successful BCG-induced protection of macaques from subsequent Mtb challenge ^43^. Type I interferon mediated restriction of mycobacterial growth in in vitro granulomas comprising collagen impregnated microspheres containing Mtb infected human peripheral blood mononuclear cells (PBMC) ^44^ also suggests that type I interferon responses benefit the host.

Our observation of reduced recruitment of macrophages to the site of disease is likely the most important factor driving increased susceptibility to *M. marinum* infection in *stat2* CRISPants. Although macrophages have been implicated in the dissemination of *M. marinum* in zebrafish larvae ^23,45^, reduction of total macrophage numbers, impaired macrophage migration to the site of mycobacterial infection or failure to control intracellular bacterial growth within macrophages have all been associated with poor outcome ^23–25,46,47^. In contrast, increasing macrophage numbers via augmentation of M-CSF signalling either pre-or post-granuloma formation, promotes resistance to *M. marinum* infection by reducing granuloma necrosis, with consequent curtailment of extracellular mycobacterial growth ^24^. The recent report that human genetically inherited CCR2 deficiency is associated with reduced pulmonary recruitment of monocytes, resulting in depleted numbers of functionally normal alveolar macrophages and increased susceptibility to mycobacteria, is also consistent with the concept that adequate numbers of macrophages at the site of infection are important for protective anti-mycobacterial immunity ^48^.

The reduction in neutrophils present at the site of *M. marinum* infection in *stat2* CRISPants may have arisen due to baseline neutropenia, defective migration or both combined. On the basis that steady state neutropenia did not affect the timing and magnitude of peak neutrophil recruitment to sterile injury, impaired migration is probably the main driver of this phenotype, which likely also contributed to increased severity of mycobacterial disease. Previous data show that reduced access of neutrophils to mycobacteria leads to increased bacterial burden in a zebrafish line in which neutrophils express mutant *cxcr4*, resulting in peripheral neutropenia and impaired neutrophil recruitment to sites of inflammation, due to their retention in haematopoietic tissue ^26,49^.

Our study has some limitations. We were not able to address the reasons for variation in type I interferon activity in our human cohort. Variation in type I interferon responses may arise due to genetically or epigenetically encoded variation in the immune response to Mtb, neutralising type I interferon auto-antibodies, burden and virulence of the mycobacterial inoculum and stochasticity in the immune response pathway following exposure ^50–54^. Our sample size is insufficient for robust detection of associations between genetic variation and the expression levels of type I interferon signalling pathway genes. Serum samples for measurement of autoantibodies to type I interferons are not available, although anti-interferon antibodies are very rare in cohorts of this age distribution ^55^. We were also unable to completely disentangle whether reduced neutrophil recruitment to the site of *M. marinum* infection was a consequence of neutropenia or impaired migration. We have not determined the mechanism by which attenuated type I interferon signalling leads to reduced myeloid cell recruitment to *M. marinum*. The expression of several chemokines is type I interferon inducible ^31,56,57^ and the CCR2-CCL2 and CXCR3-CXCL11 chemokine axes are known to be involved in macrophage recruitment to *M. marinum* ^45,58^. Future studies are required to test the hypothesis that *stat2* mutagenesis disrupts chemokine-mediated cellular recruitment to the site of mycobacterial disease. Nonetheless, our work provides new insights into type I interferon mediated host protection against mycobacterial infection.

Our findings have potential implications for risk stratification of individuals in whom any of the above mechanisms predisposing to low type I interferon activity are identified. In active TB, such individuals could be targeted for longer or more intensive drug regimens, or adjunctive type I interferon therapy delivered to the respiratory tract, to mitigate against severe disease. Early intervention with type I interferon treatment, before destruction of the lung parenchyma, is likely to be crucial to augment macrophage containment of Mtb, but timing and duration of therapy will require evaluation in relatively large-scale experimental medicine trials.

## Methods

### Ethics statement

The study was approved by the London Bloomsbury Research Ethics Committee (16/LO/0776). Written informed consent was obtained from all participants.

### Study design

We aimed to identify human immune pathways that correlate with severity of TB disease, using transcriptional profiling of biopsies from the TST as a surrogate for immune responses to Mtb in the lung, and peripheral blood samples. We tested causation for observed associations by genetic manipulation of identified pathways in the zebrafish larval mycobacterial infection model. To encompass a broad spectrum of disease severity we recruited 51 adults (>18 years) from four London hospitals, newly diagnosed with smear or culture positive pulmonary TB, who were within four weeks of starting treatment. This is in keeping with the median sample size of previous studies which have successfully demonstrated a relationship between severity of TB disease and immunological parameters ^7,59–62^. Individuals with coincident human immunodeficiency virus infection, malignancy, taking immunomodulatory therapy, a history of immunization within the preceding two weeks or previous keloid formation, or unable to give informed consent were excluded. Study participants underwent blood collection into Tempus tubes before intradermal injection of 0.1 ml of two units of tuberculin (AJ Vaccines) or saline in the volar aspect of the forearm as previously described ^8–10^. At 48 hours clinical induration at the injection site was measured, immediately prior to sampling by 3mm punch biopsy as previously described ^8,9^. Additional saline samples from individuals with cured or latent TB, BCG vaccine recipients and healthy volunteers have been described previously ^10^. Control blood samples from healthy individuals have been described previously ^13^.

### RNA purification from blood, skin biopsy and zebrafish embryo samples

Total blood RNA was extracted using the Tempus Spin RNA Isolation Kit (Ambion; Life Technologies). Globin mRNA was removed using the GlobinClear kit (Life Technologies). Skin biopsies were homogenized as previously described ^10^. Pooled zebrafish embryos (25-30 per experimental condition) collected in Qiazol (Qiagen) were homogenized in CK14 lysing kit tubes (Bertin Instruments) using a Precellys Evolution homogenizer (Bertin Instruments) for 10-20 seconds at 5500 rpm. Total RNA from skin biopsies and zebrafish samples was purified using the RNEasy mini and micro kits (Qiagen), respectively, according to the manufacturer’s instructions. Genomic DNA was removed using the TURBO DNA-free kit (ThermoFisher).

### Transcriptional profiling of blood, skin biopsy and zebrafish larval samples

Genome-wide mRNA sequencing was performed as previously described ^10,63^. Complementary DNA (cDNA) libraries were prepared using the KAPA HyperPrep kit (Roche). Sequencing was performed in paired-end mode with the NextSeq 500 High Output 75 Cycle Kit (Illumina) using the NextSeq500 system (Illumina) according to the manufacturer’s instructions, generating a median of 24 million (range 14.7-32.4 million) 41 base pair reads per human blood sample, 22 million (range 12.3-27.9 million) reads per human TST sample and 16.8 million (range 15.1-18.1 million) reads per zebrafish sample. Transcript alignment and quantitation against the Ensembl GRCh38 human genome assembly release 100 or GRCz11 zebrafish genome assembly release 104 was performed using Kallisto ^64^. Transcript-level output counts and transcripts per million (TPM) values were summed on gene level and annotated with Ensembl gene ID, gene name, and gene biotype using the R/Bioconductor packages tximport and BiomaRt ^65,66^. Downstream analysis was performed using R 3.6.3 on log2 transformed TPM values for protein-coding genes only. 0.001 was added to all TPM values prior to log2 transformation. To determine the need for and effect of batch correction in the human data, principal component analysis (PCA) was performed using the prcomp R function on the integrated list of genes with the 10% lowest variance in expression for each library preparation date. This revealed separation of three TST samples in PC1, likely accounted for by lower RNA concentrations or RIN values compared to the rest of the dataset (Figure S1A). After batch correction performed using the ComBat function of the sva R package ^67^, allocating samples with PC1 score < and >0 to separate batches, this was no longer evident (Figure S1B).

### Radiographic quantitation of TB severity

Disease severity was evaluated by estimating the extent of radiographic abnormalities and whether cavitation was present on chest x-ray at the time of diagnosis, as previously described ^10,11^.

### Outlier analysis

The BioBase Bioconductor R package was used to perform outlier analysis based on the z-score outlier detection (ZODET) algorithm ^12^, to identify transcripts in each TB patient blood sample whose expression was significantly higher than the mean expression in control blood samples from healthy volunteers ^13^ and transcripts in each TST sample whose expression was significantly higher than the mean expression in control saline samples (>2SD, p value <0.05), with a fold difference filter of >1 log2. Chi-squared tests implemented in R were then used to identify outlier genes statistically enriched in blood samples from TB patients compared to healthy individuals and TST samples compared to saline samples, using a p value <0.05. To avoid type 2 error no correction for multiple testing was used in this analysis. In instances where multiple Ensembl gene IDs were associated with a single gene symbol, duplicate gene symbols were removed to retain the replicate with the highest expression for each sample.

### Pathway enrichment analysis

The biological pathways represented by the entire blood and TST transcriptomes and genes in each transcriptome positively and negatively correlated with radiographic severity score (Spearman rank correlation p value <0.05, without correction for multiple testing, to prevent type 2 error) were identified by Reactome pathway enrichment analysis using XGR as previously described ^68,69^. For visualization, 15 pathway groups were identified by hierarchical clustering of Jaccard indices to quantify similarity between the gene compositions of each pathway. For each group the pathway term with the largest number of annotated genes was then selected as representative of enriched biology.

### Upstream regulator analysis

Peripheral blood and TST transcriptome genes whose expression was significantly correlated with radiographic severity of TB disease were identified by Spearman rank correlation, p value <0.05, without correction for multiple testing, to prevent type 2 error. Genes positively and negatively correlated with disease severity were separately subjected to Ingenuity Pathway Analysis (Qiagen) to identify upstream molecules responsible for their transcriptional regulation. This analysis was restricted to molecules annotated with the following functions: cytokine, transmembrane receptor, kinase and transcriptional regulator, representing the canonical components of pathways which mediate transcriptional reprogramming in immune responses. Enriched molecules with an adjusted p value <0.05 were considered statistically significant.

### Upstream regulator functional modules

Blood and TST gene modules representing the functional activity of their upstream molecule were identified as previously described ^13^. In brief, we calculated the average correlation coefficient for pairwise correlations of the expression levels of the target genes associated with each predicted upstream regulator. We compared these to the distribution of average correlation coefficients obtained from 100 iterations of randomly selecting equivalent sized groups of genes from the blood or TST transcriptome. Only target gene modules with average correlation coefficients higher than the mean of the distribution of equivalent sized randomly selected groups by ≥2 SD (Z score ≥2) with false discovery rate <0.05, were retained. We visualised these upstream regulator modules as network diagrams using the Force Atlas two algorithm in Gephi v0.9.2 ^70^, depicting all statistically over-represented molecules (false discovery rate <0.05) predicted to be upstream of ≥4 target genes.

### Transcriptional modules

Interferon inducible transcriptional module gene lists are provided in Table S1. Independently derived human type I and type II interferon inducible gene modules were described previously ^9,10,13^. Two further modules were derived to distinguish type I or type II interferon inducible gene expression using published transcriptional data for primary human keratinocytes stimulated with a selection of cytokines (Gene Expression Omnibus accession: GSE36287) ^71^. Stimulus-specific modules for type I interferon (interferon alpha) and type II interferon (interferon gamma) were derived by selecting genes overexpressed >4-fold in the cognate cytokine condition relative to all non-cognate conditions combined (paired t test with α of p<0.05 without multiple testing correction), excluding genes also induced by each non-cognate cytokine condition, using the same criteria. The babelgene R package (https://igordot.github.io/babelgene/) was used to convert human gene symbols to their respective zebrafish orthologues to generate the zebrafish Stat1 module, excluding zebrafish genes reported to be an orthologue of more than one human gene. The zebrafish type I interferon inducible gene module comprises 360 genes whose expression was significantly induced (≥2 fold, adjusted p value <0.05) six hours after intravenous injection of three day post-fertilisation (dpf) larvae with 1 nl of recombinant interferon phi 1 protein (1 mg/ml) as previously described ^18^.

### Venn diagrams

Area-proportional Venn diagrams visualizing the overlap between constituent genes in transcriptional modules were generated using BioVenn ^72^.

### Zebrafish husbandry

Zebrafish were raised and maintained at 28.5°C following a 14/10 light/dark cycle according to standard protocols ^73^ in University College London (UCL) and Neuro-PSI Zebrafish Facilities. All work was approved by the British Home Office (Project License PP8780756) and the French Ministry for Research and Higher Education (Project License 39238). All experiments were performed in accordance with the relevant guidelines and regulations, on larvae up to five days post-fertilisation, before they are protected under the Animals (Scientific Procedures) Act. Adult zebrafish were spawned to collect embryos for experiments. Table S2 lists the lines used in this study ^22,28,29^. Embryos were maintained at 28.5°C in egg water containing 60 μg/ml Tropic Marin Sea Salt (Norwood Aquarium) and anaesthetised with egg water containing 200 µg/ml buffered tricaine (3-aminobenzoic acid ethyl ester) (Sigma-Aldrich) during injection, tail fin transection and imaging. Larvae used for in vivo imaging were maintained in egg water supplemented at 24 hours post-fertilisation (hpf) with 0.003% PTU (1-phenyl-2-thiourea) (Sigma-Aldrich) to inhibit melanisation.

### CRISPant Generation

To maximise the likelihood of loss of functional protein we employed a previously described triple-exon targeting approach ^74^, with the modification that Cas9 protein was not diluted before use. CRISPR RNA (crRNA) (Alt-R CRISPR-Cas9 crRNA), tracrRNA (Alt-R CRISPR-Cas9 tracrRNA) and recombinant Cas9 protein (Alt-R Streptococcus pyogenes Cas9 Nuclease V3) were obtained from Integrated DNA Technologies (IDT). Three pre-designed crRNAs targeting distinct exons were selected for the *stat2* gene, prioritizing those that targeted early, asymmetric (not divisible by three) exons common to both protein-coding transcripts, with low off-target and high on-target scores and in a genomic position which allowed design of primers flanking the target site to generate a ∼200 bp amplicon, for verification of mutagenesis by next generation sequencing (MiSeq). crRNA sequences and genomic locations are provided in Table S3. To generate gRNAs equal volumes of crRNA and tracrRNA diluted to 61 μM in Duplex buffer (IDT) were annealed at 95°C for 5 minutes. Equal volumes of gRNA (61 μM) and Cas9 protein (61 μM) were incubated at 37°C for 5 minutes to assemble separate 30.5 μM ribonucleoproteins (RNPs) for each target. Equal volumes of the three RNPs were pooled, prior to injection giving a final concentration of 10.2 μM for each. Negative control “scrambled” crRNAs computationally designed to be non-targeting (Alt-R CRISPRCas9 Negative Control crRNA #1, #2, #3) were prepared and pooled as described above. 0.5 nl of pooled *stat2* RNPs or scrambled RNPs was injected into the yolk sac of single-cell stage wild type or transgenic zebrafish embryos before cell inflation as previously described ^74^. After injections embryos were transferred into sterile petri dishes with fresh egg water and maintained at 28.5°C to the appropriate age for experiments, as described below.

### Interferon phi 1 stimulation

Three dpf *stat2* CRISPant *Tg(mxa:mCherry)* larvae and scrambled RNP injected siblings were injected in the coelomic cavity with 1 nl of recombinant zebrafish interferon phi 1 protein (1.25 mg/ml), produced as previously described ^75^. *stat2* and control CRISPants were shuffled so that injections were performed blindly. Bovine serum albumin (BSA) (1.25 mg/ml) was injected as a control. Interferon phi 1 and BSA solutions were supplemented with 0.2% phenol red (Sigma-Aldrich) to visualize the inoculum. Induction of Mxa reporter expression was determined by fluorescence microscopy 24 hours post-injection.

### M. marinum infection

*M. marinum* M strain expressing the pmsp12 mWasabi ^76^ or psMT3 mCherry plasmid ^77^ was cultured using Middlebrook 7H10 agar (Becton Dickinson and Company) supplemented with 0.5% oleic acid/albumin/dextrose/catalase (OADC) (Becton Dickinson and Company), 0.5% glycerol (Sigma-Aldrich) and hygromycin (50 μg/ml) (Fisher Scientific). To generate bacterial suspensions for injection, *M. marinum* from agar plates was cultured statically at 28.5°C for 24-36 hours in 10 ml Middlebrook 7H9 broth (Becton Dickinson and Company) supplemented with 10% albumin/dextrose/catalase (ADC) (Becton Dickinson and Company) and hygromycin (50 μg/ml) (Fisher Scientific). Bacteria were harvested at mid-log growth (optical density (OD)600nm 0.7–1), (OD)600nm 1 representing 10^8^ colony forming units (cfu)/ml, washed three times in phosphate buffered saline (PBS) (Gibco) and resuspended in 2% polyvinylpyrrolidone (PVP)40 (Sigma-Aldrich)/PBS containing 10% phenol red (Sigma-Aldrich) to aid visualisation of injections ^78^.

Zebrafish embryos were manually dechorionated using jeweller’s forceps (Dumont #5, World Precision Instruments) prior to bacterial infection. To generate systemic infection, 400 cfu *M. marinum* in a volume of 1 nl were microinjected into the caudal vein of embryos staged at 28-30 hpf. Four days post-infection, embryos were either subjected to fluorescence microscopy to measure disease severity or 25-30 per condition pooled and harvested in Qiazol (Qiagen) for evaluation of immune responses by RNAseq. To generate localised infection, 200 cfu *M. marinum* in a volume of 1 nl were injected into the hindbrain ventricle (HBV) of two dpf zebrafish larvae. Cellular recruitment was evaluated by fluorescence microscopy at 6-18 hours post-infection.

### Tailfin transection

Tailfin transections were performed as described previously ^29,79^. Briefly, live anaesthetised three dpf zebrafish larvae were transferred with a 1 ml Pasteur pipette (Scientific Laboratory Supplies) to a sterile petri dish containing 2% agarose (Bioline). Transections of the caudal fin were performed using a sterile microscalpel (World Precision Instruments) by applying steady downward pressure to create the incision at the boundary with the notochord. Following injury, larvae were rinsed in egg water and maintained in individual wells of a 96-well tissue culture plate (TPP) at 28.5°C until live imaging to quantify neutrophil or macrophage recruitment to the site of injury at 1, 6 and 24 hours-post-wound (hpw).

### Fluorescence microscopy

All fluorescence microscopy was performed on live, anasethised larvae. For quantitation of bacterial burden and dissemination four days post-infection, steady state neutrophil numbers at two dpf and cellular recruitment at 2-4 dpf, larvae were imaged using an M205FA stereofluorescence microscope (Leica) with a 1x objective. Larvae were imaged on a flat 1% agarose plate for quantitation of severity of mycobacterial infection and steady state neutrophil numbers and a 96-well tissue culture plate (TPP) in which the wells were coated with 1% agarose, for cellular recruitment to tailfin transection. Micro-wells were created in 1% agarose using a Stampwell Larvae 1 device (Idylle) for optimal positioning of 2-3 dpf larvae for imaging of cellular recruitment to hindbrain ventricle *M. marinum* infection. Brightfield and fluorescence images were captured using a DFC365 FX camera (Leica) and exported as 16-bit Tagged Image Format (TIF) files for analysis. For quantitation of steady state macrophage numbers three dpf larvae were imaged in 96-well alignment plates (Funakoshi) containing 75 μl egg water per well using a high-content wide-field fluorescence microscope (Hermes, IDEA Bio-Medical) with a 4x objective, capturing five z planes 50.6 μm apart in four contiguous fields of view per embryo. We created an ImageJ macro to semi-automate generation of single maximum intensity projection z-stack montages from the Hermes images, which were saved as 16-bit TIF files for analysis (Table S4). Mxa reporter expression was recorded with an EVOS M7000 microscope (ThermoFisher) equipped with a 2x/NA0.06 objective and Texas Red fluorescence cube. Larvae were positioned laterally in micro-wells created in 2% agarose with an in-house-printed stamp. A single fluorescence plane was recorded as the depth of field is sufficient to capture the entire larva. Transmitted light images were also recorded and both were exported as 16-bit TIF files.

### Image analysis

To quantify bacterial burden, number of bacterial foci and spatial distribution of bacteria, images were analysed using custom QuantiFish software, previously developed in our group, to enable rapid quantitation of fluorescent foci in zebrafish larvae ^80^. Mxa reporter fluorescence was measured with ImageJ. Transmitted and fluorescence images were stacked. Images were slightly rotated to horizontality where necessary. A 600x100 pixel rectangular region of interest was defined on the transmitted light image to encompass the entire gut and liver areas. Mean fluorescence intensity was then measured on the fluorescence image. For quantitation of total cell number and recruitment of fluorescent immune cells to the site of tailfin transection we developed a new custom Python script which enables quantitation of integrated fluorescence within concentric zones (known as Sholl circles) ^81^ (Figure S9). To achieve this, the script leverages the OpenCV, NumPy ^82^, Matplotlib, SciPy (http://www.scipy.org), scikit-image ^83^ and pandas Python libraries to facilitate image processing and data extraction (Table S5). First the user is prompted to select a folder containing images for analysis, as well as the file extensions corresponding to the brightfield and fluorescence images, which are essential for the subsequent steps. The script then iterates through each pair of brightfield and fluorescent images prompting the user to define two points on the brightfield image, which serve as the start and end positions between which to create concentric circular regions ^81^ on the corresponding fluorescence image. The script employs the image processing techniques described in Table S5 to quantify integrated fluorescence of the entire image and that within each region, as surrogates for the total number of immune cells and the number at each spatial location, respectively and generates output files containing this information. Integrated fluorescence within the first zone (the wound site) and distance from the site of tailfin transection of the zone in which the maximal integrated fluorescence (a surrogate for the highest cell numbers) was detected (identified manually) were used as measures of immune cell recruitment for each zebrafish larva. The number of cells recruited to the site of hindbrain *M. marinum* infection was quantified by manual counting.

### Genotyping by next generation sequencing

#### Genomic DNA extraction

Genomic DNA was extracted from anasethetised individual zebrafish larvae using the modified hot sodium hydroxide and Tris (HotSHOT) method as previously described ^74,84^. Briefly, larvae in 96-well plates were incubated in 50 μl base solution (25 mM KOH, 0.2 mM EDTA in water) at 95°C for 30 min then cooled to room temperature, before addition of 50 μl neutralization solution (40 mM Tris-HCL in water).

#### PCR

Extracted DNA (2 μl per 50 μl reaction) was subjected to PCR using Phusion High-Fidelity DNA Polymerase (New England Biolabs) (98°C for 30 seconds (s), once only, 40 cycles of 98°C for 10 s, 59°C for 30 s, 72°C for 20 s, 72°C for 10 minutes, once only). Sequences and genomic positions of the primers used are provided in Table S3. Successful amplification of a single product of approximately 300 bp was verified by 2% agarose (Bioline) Tris-borate-EDTA (TBE) containing Redsafe (Chembio) gel electrophoresis. PCR products were purified using ExoSAP-IT Express (ThermoFisher), concentration of purified DNA determined by Nanodrop spectrophotometer (ThermoFisher) and samples diluted to 15-25 ng/μl for sequencing.

#### Next generation sequencing and data analysis

Sequencing was performed by the UCL Fish Genomic Service using the MiSeq Reagent Kit v2 (300-cycles) (Illumina) and the MiSeq system (Illumina) in paired end mode, according to the manufacturer’s instructions. Sequencing data were received as two FASTQ files (forward and reverse) for each sample. Visual sequence analysis was performed using Geneious 10.2.6 (https://www.geneious.com) for initial verification of successful genome editing close to the PAM site. Objective quantitation of the proportion of reads mutated and predicted to be frameshifted was achieved using the ampliCan R package ^85^, which utilizes the FASTQ files and a metadata file providing the amplicon, protospacer adjacent motif (PAM) and primer sequences (Table S3) and sample information, as previously described ^74^. AmpliCan was run with default parameters, with the exception that min_freq = 0.005 (mutations at a frequency below this threshold were considered a sequencing error) and average_quality = 25. Mutations present in uninjected or scrambled RNP injected control samples were not included as Cas9 induced mutations.

### Statistical analysis

Statistical analysis was performed using GraphPad Prism 9.3.1 for Windows (GraphPad Software) and R3.6.3. Chi-squared tests were used to identify outlier genes statistically enriched in transcriptomic data from blood samples from people with TB compared to healthy individuals and TST samples compared to saline samples, with no correction for multiple testing to avoid type 2 error. Spearman rank correlation was used to determine the strength of relationship between blood and TST transcriptome gene expression and TB disease severity (two-tailed), transcript levels of upstream regulator target gene modules, blood and TST random gene modules (one-tailed, positive correlation only) and mean expression of transcriptional modules in blood and TST transcriptomic data with severity of TB disease (two-tailed). The Benjamini-Hochberg correction method for multiple comparisons was used to identify upstream regulator target gene modules with average correlation coefficients significantly higher than the mean of the distribution of equivalent sized randomly selected gene modules by ≥2 SD (Z score ≥2) with false discovery rate <0.05. Survival of *stat2* CRISPant and control zebrafish larvae was compared using a log-rank (Mantel-Cox) test; a p value <0.05 was considered significant. On scatter plots horizontal lines represent median values and error bars indicate the interquartile range. On box and whisker plots horizontal lines represent median values, box limits indicate the interquartile range and whiskers extend between the 5th and 95th percentiles of the data. Two-tailed Mann-Whitney tests were used for two group comparisons of unpaired data; a p value <0.05 was considered significant. Where p values are not presented on graphs, * denotes p<0.05.

## Data Availability

Human TST RNAseq data and peripheral blood RNAseq data from healthy individuals are already available in the ArrayExpress repository under accession numbers E-MTAB-6816 and E-MTAB-10022, respectively. Human peripheral blood data from TB patients and zebrafish RNAseq data will be made available in ArrayExpress at the time of peer reviewed publication of the manuscript. Source data for figure panels with fewer than 20 data points are available in Data S1.

https://www.ebi.ac.uk/biostudies/arrayexpress/studies/E-MTAB-6816

https://www.ebi.ac.uk/biostudies/arrayexpress/studies/E-MTAB-10022

## Acknowledgements

We thank Michelle Berin and Victoria Dean (Clinical Research Nurses, UCL Division of Infection and Immunity) and Mylah Ramirez (Research Nurse, Barts Health NHS Trust) for support with research governance, participant recruitment and sample collection. We thank the UCL Zebrafish Facility staff, particularly Heather Callaway, Jenna Hakkesteeg, Karen Dunford, Carole Wilson and Elise Hitchcock for maintenance of the zebrafish lines used in this study, especially during the first wave of the COVID-19 pandemic. We thank UCL Genomics for library preparation and sequencing of samples for transcriptional profiling (www.ucl.ac.uk/child-health/research/genetics-and-genomic-medicine/ucl-genomics). We thank Alex Lubin and Katie-Jo Miller for next generation sequencing of zebrafish samples for genotyping. This work has benefited from the facilities and expertise of CELPHEDIA-TEFOR, UAR2010 TEFOR Paris-Saclay (TEFOR Infrastructure - Investissement d’avenir - ANR-II-INBS-0014) and from the Recombinant Protein Platform of the Institut Pasteur, Paris.

GST is supported by a UK MRC Clinician Scientist Fellowship (MR/N007727/1). MN is supported by the Wellcome Trust (207511/Z/17/Z). PME is supported by a Sir Henry Dale Fellowship jointly funded by the Wellcome Trust and the Royal Society (105570/Z/14/Z). J-PL is supported by Fondation pour la Recherche Medicale (grant EQU202203014646). MN and GST are also supported by NIHR Biomedical Research Centre Funding to University College Hospitals NHS Foundation Trust and University College London.

## Author contributions

Conceived the study: GST, MN

Collected the data: ASS, BSMDDE, JR, AC, ET, GP, J-PL, GST

Provided resources or analysis tools: ASS, BSMDDE, JR, CTT, MCIL, HK, GP, PME, EMP, J-PL, MN, GST

Performed the data analysis / interpretation: ASS, BSMDDE, JR, CTT, GP, PME, EMP, J-PL, MN, GST

Prepared the manuscript draft: GST, ASS, BSMDDE

All authors reviewed and contributed to the final manuscript.

## Code availability

The source code and manual for QuantiFish software used for analysis of bacterial burden and dissemination in zebrafish larvae are available at https://github.com/DavidStirling/QuantiFish. The ImageJ macro used to generate montages from the Hermes microscope images and source code and manual for the Python script used for analysis of steady state cell numbers and cellular recruitment in zebrafish are available at https://github.com/andreaszydloshein/FIJI-Image-to-Stack and https://github.com/andreaszydloshein/Zytometer, respectively.

