## Supplemental information for "Type I interferon responses contribute to immune protection against mycobacterial infection"

### Title

*Corresponding author

### Corresponding author

### LIST OF SUPPLEMENTAL INFORMATION

Figure S1. Batch correction of tuberculin skin test (TST) transcriptomic data from people with active tuberculosis (TB).

Figure S2. Interindividual variation and pathway enrichment analysis of peripheral blood transcriptomic data from people with active TB.

Figure S3. Identification of upstream regulator target gene modules in the TB peripheral blood transcriptome.

Figure S4. Interindividual variation and pathway enrichment within the TB TST transcriptome.

Figure S5. Identification of biological pathways and upstream regulator target gene modules among TB TST transcriptome genes that correlate with radiographic disease severity.

Figure S6. Relationship between the expression of the entire TST transcriptomic signature and interferon inducible gene modules with TB disease severity.

Figure S7. Schematic representation of zebrafish experiments.

Figure S8. *stat2* mutagenesis blocks type I interferon signalling in zebrafish.

Figure S9. Quantitation of cellular recruitment to the site of sterile tailfin transection.

Table S1. Interferon response modules.

Table S2. Zebrafish lines.

Table S3. *stat2* crRNAs, primers and amplicon sequences.

Table S4. ImageJ macro description.

Table S5. Python script workflow and programming.

Data S1. Collated original data for figure panels with fewer than 20 data points.

This file includes Figures S1-S9 and Tables S2, S4 and S5.

Tables S1 and S3 and Data S1 are provided as separate files.

### Supplementary figures

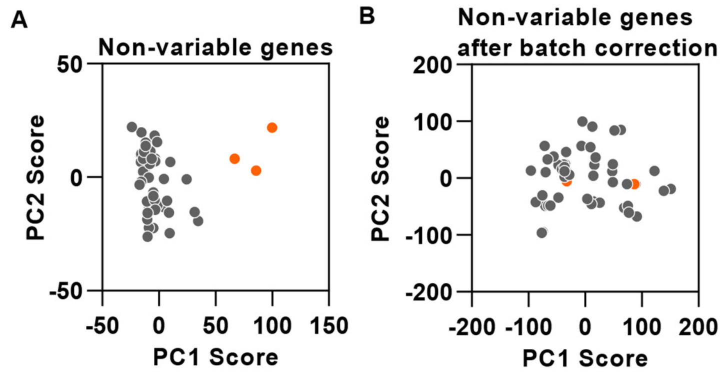


**Figure S1. Batch correction of tuberculin skin test (TST) transcriptomic data from people with active tuberculosis (TB), related to Table 1, Figure 2 and Transcriptional profiling of blood and skin biopsy samples in Methods.** Principal component analysis of the integrated list of the 10% genes in TST transcriptomic data with least interindividual variance in expression, from each library preparation run before **(A)** and after **(B)** batch correction. PC = Principal component.

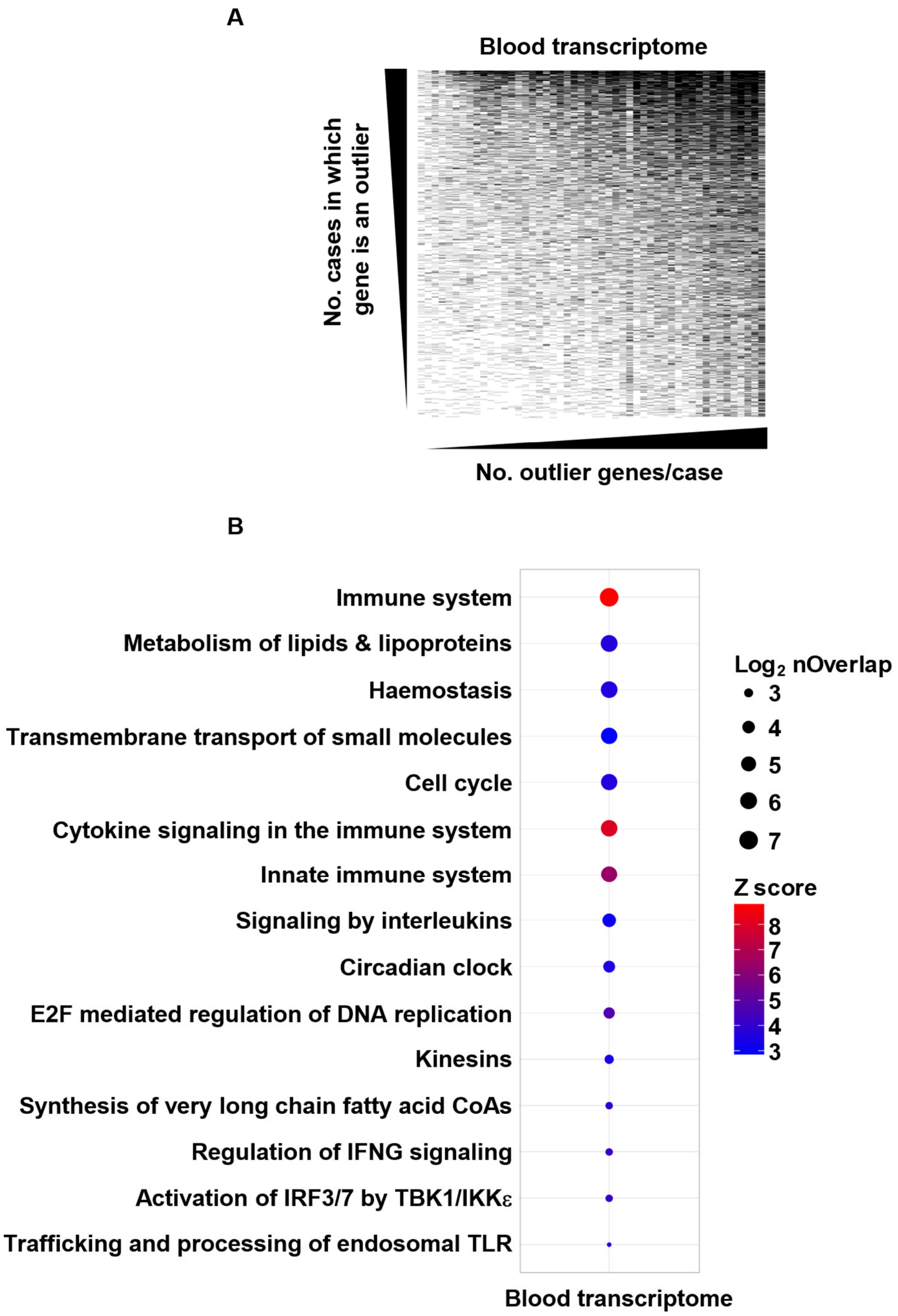


**Figure S2. Interindividual variation and pathway enrichment analysis of peripheral blood transcriptomic data from people with active TB, related to Figure 1. (A)** Representation of the integrated list of 2620 outlier transcripts derived from the entire cohort, which comprise the TB peripheral blood transcriptome. The matrix depicts blood transcriptome genes (rows) for each study subject (columns); outlier transcripts are indicated in black. **(B)** Reactome pathway enrichment within the TB peripheral blood transcriptome.

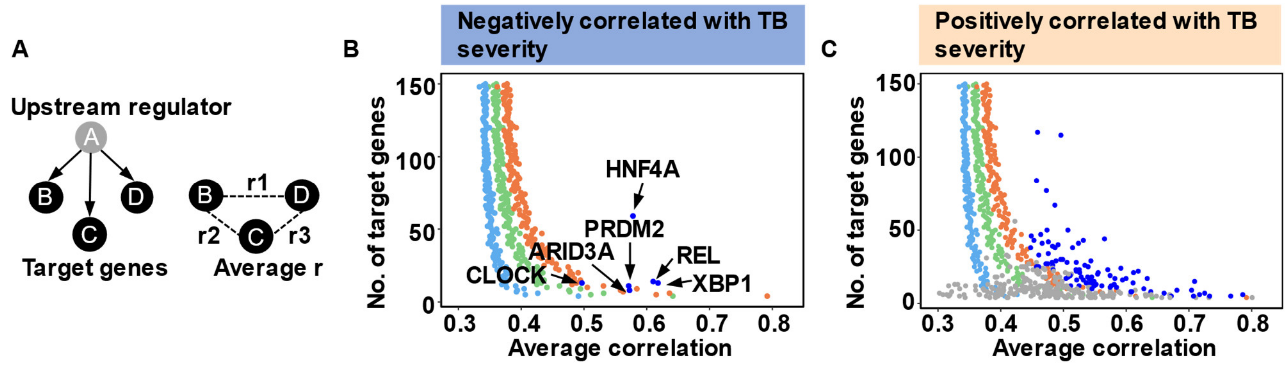


**Figure S3. Identification of upstream regulator target gene modules in the TB peripheral blood transcriptome, related to Figure 1. (A)** Schematic of a putative functional gene network (target genes B,C,D) regulated by an upstream molecule predicted to determine its expression (gene A). Correlation coefficients for the expression levels of its component genes are indicated (r1, r2, r3). **(B, C)** Z-scores derived from the distribution of average correlation coefficients obtained from 100 iterations of randomly selecting groups of genes from the TB blood transcriptome are shown in light blue (z-score=1), green (z-score=2) and orange (z-score=3). Average correlation coefficients of upstream regulator target gene modules identified within blood transcriptome genes negatively correlated **(B)** or positively correlated **(C)** with disease severity are shown in dark blue (z-score ≥2, FDR ≤0.05) and grey (z-score ≤2 +/- FDR ≥0.05) compared to equivalent sized random gene modules. Among genes negatively correlated with TB severity, target gene modules that show significantly greater co-correlated expression compared to random gene modules are annotated by their predicted upstream regulator **(B)**.

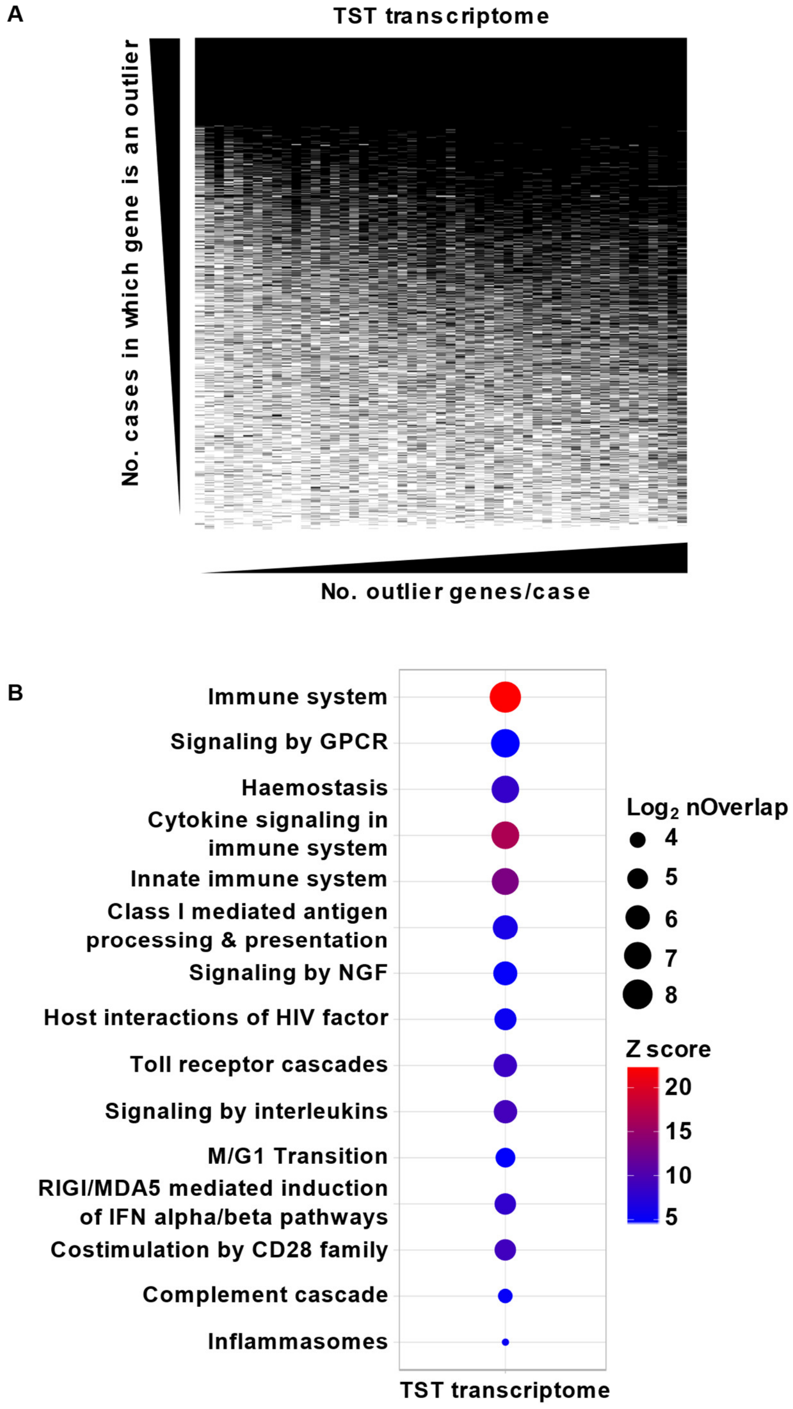


**Figure S4. Interindividual variation and pathway enrichment within the TB TST transcriptome, related to Figure 2. (A)** Matrix representing the integrated list of 3222 outlier transcripts derived from the entire cohort, which comprise the TB TST transcriptome. Rows represent each gene and columns each TST sample; outlier transcripts are indicated in black. **(B)** Reactome pathway enrichment within the TB TST transcriptome.

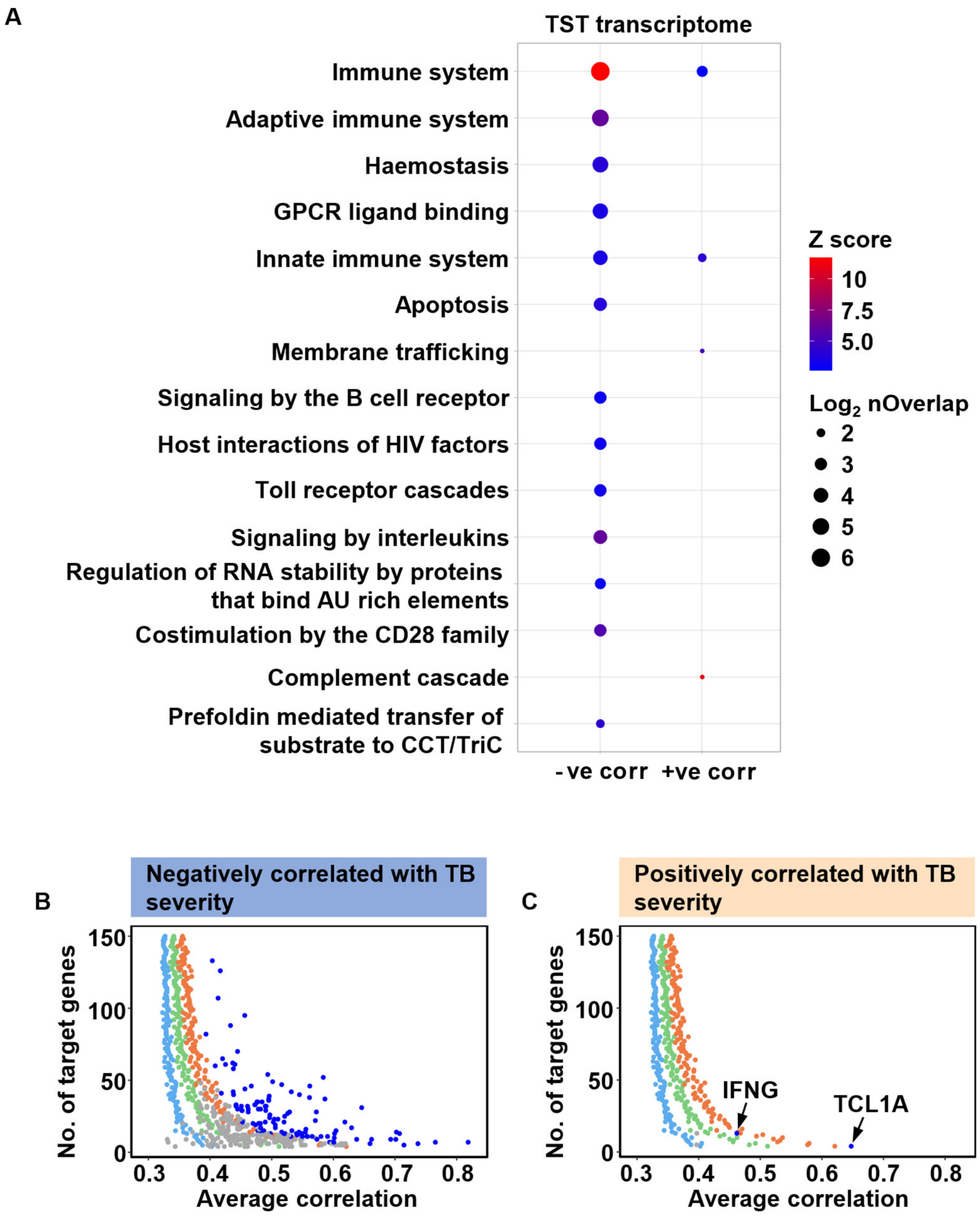


**Figure S5. Identification of biological pathways and upstream regulator target gene modules among TB TST transcriptome genes that correlate with radiographic disease severity, related to Figure 2. (A)** Reactome pathway enrichment among TST transcriptome genes whose expression is statistically significantly correlated with radiographic TB disease severity; corr = correlated. **(B, C)** Z-scores derived from the distribution of average correlation coefficients obtained from 100 iterations of randomly selecting groups of genes from the TB TST transcriptome are shown in light blue (z-score=1), green (z-score=2) and orange (z-score=3). Average correlation coefficients of upstream regulator target gene modules identified within TST transcriptome genes negatively correlated **(B)** or positively correlated **(C)** with TB disease severity are shown in dark blue (z-score ≥2, FDR ≤0.05) and grey (z-score ≤2 +/- FDR ≥0.05) compared to equivalent sized random gene modules. Among genes positively correlated with TB severity, target gene modules that show significantly greater co-correlated expression compared to random gene modules are annotated by their associated upstream regulator **(C)**.

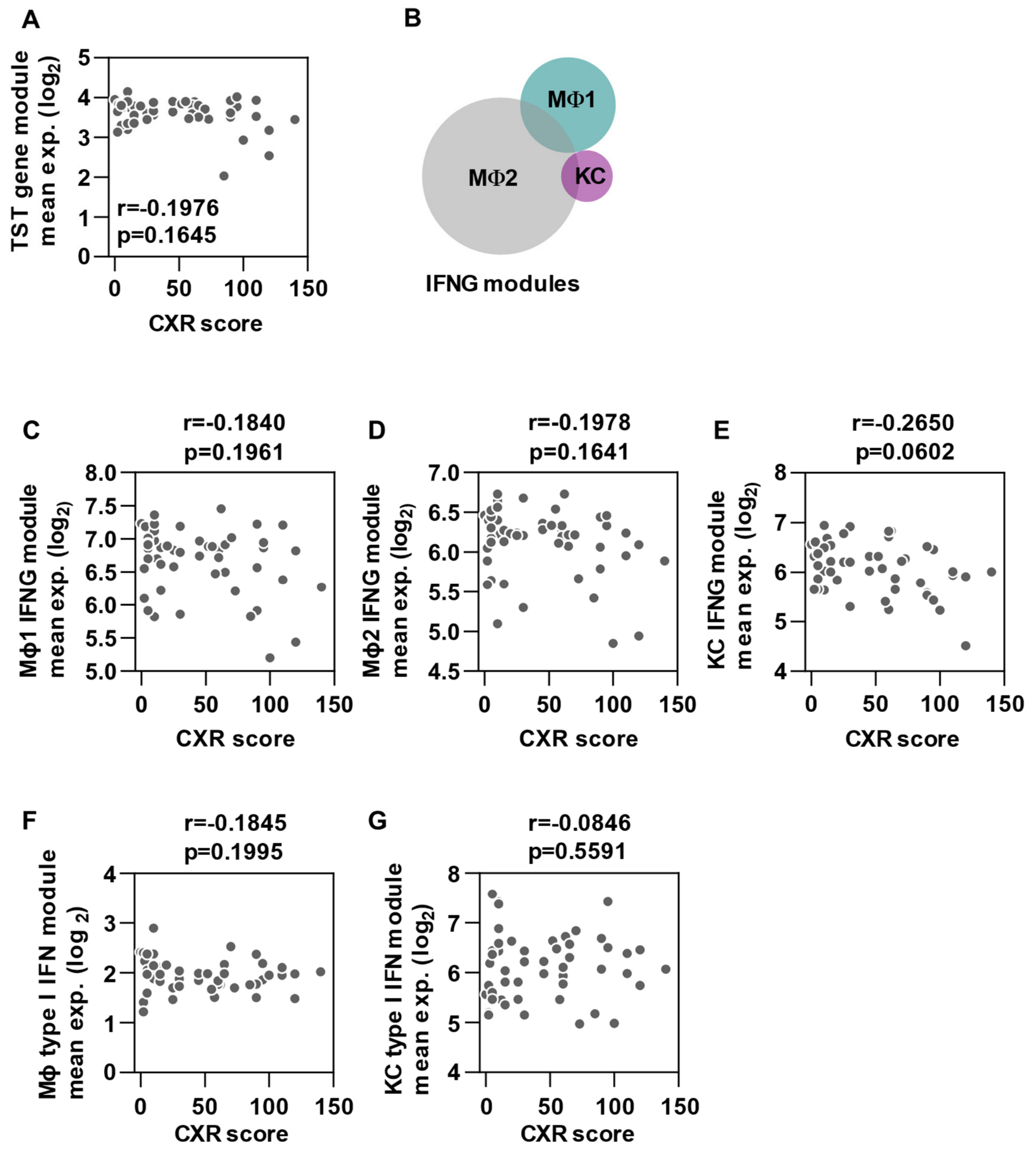


**Figure S6. Relationship between the expression of the entire TST transcriptomic signature and interferon inducible gene modules with TB disease severity, related to Figures 1 and 2.** **(A-E)** Average expression of the 3222 gene TST transcriptome **(A)** and three largely non-overlapping interferon gamma inducible gene modules derived from independent experimental data, in TST transcriptomic data **(B-E)** is not statistically significantly correlated with radiographic TB severity. **(F-G)** Average expression of two distinct independently derived gene modules specific for type I IFN activity in TB peripheral blood transcriptomic data does not show statistically significant correlation with disease severity. r values and p values were derived from two-tailed Spearman rank correlations. CXR = Chest x-ray, MΦ = Macrophage, KC = Keratinocyte.

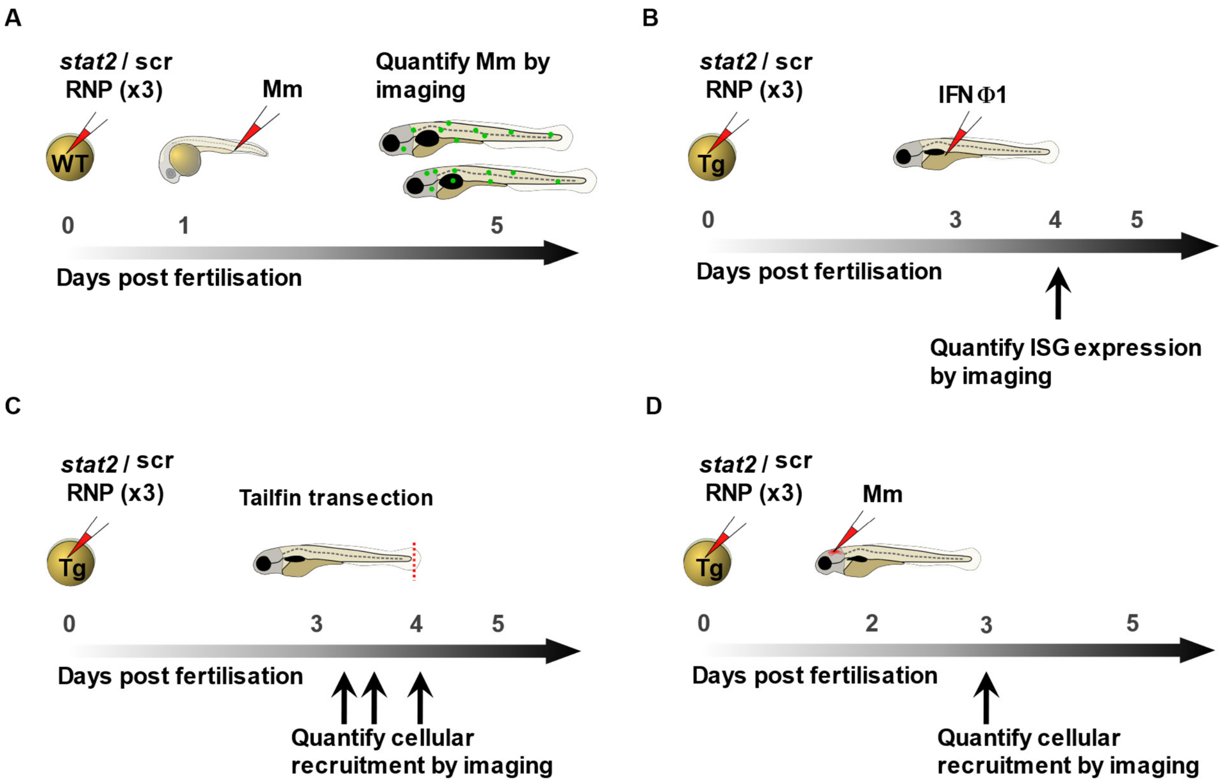


**Figure S7. Schematic representation of zebrafish experiments, related to Figures 3-7. (A)** Three guide RNA / Cas9 ribonucleoproteins (RNPs) targeting distinct exons of *stat2* are injected into the yolk sac of wild type (WT) early one cell stage embryos to generate *stat2* CRISPants. Embryos from the same clutch are injected with three negative control “scrambled” RNPs (scr) which have no genomic target. 28-30 hour post-fertilisation control and *stat2* CRISPant embryos are intravenously infected with 400 colony forming units (cfu) of fluorescent *M. marinum* (Mm). Bacterial burden and dissemination are evaluated four days after inoculation by quantitative fluorescence microscopy. **(B-D)** *Tg(mxa:mCherry), Tg(mpeg1:mCherry)* and *Tg(mpx:eGFP)* embryos (Tg) are injected with three *stat2* RNPs or scr RNPs as described in **(A)** to generate CRISPant embryos with fluorescent Mxa protein expression, macrophages or neutrophils, respectively. **(B)** Three day post-fertilisation (dpf) *stat2* CRISPant and control transgenic larvae are injected via the coelomic cavity with 1 nl of recombinant interferon phi 1 protein (1.25 mg/ml). Induced expression of the interferon stimulated gene (ISG) Mxa is measured by fluorescence microscopy at 24 hours. **(C)** A small portion of the tail fin is removed using a sterile microscalpel from three dpf *stat2* CRISPant and control transgenic embryos. Cellular recruitment to the site of sterile injury is assessed at 1, 6 and 24 hours post-wound, using quantitative fluorescence microscopy. **(D)** Two dpf *stat2* CRISPant and control transgenic embryos are infected with 200 cfu of *M. marinum* by injection into the hindbrain ventricle. Recruitment to the site of localized *M. marinum* infection is evaluated by fluorescence microscopy at 18 hours for macrophages and 6 hours for neutrophils.

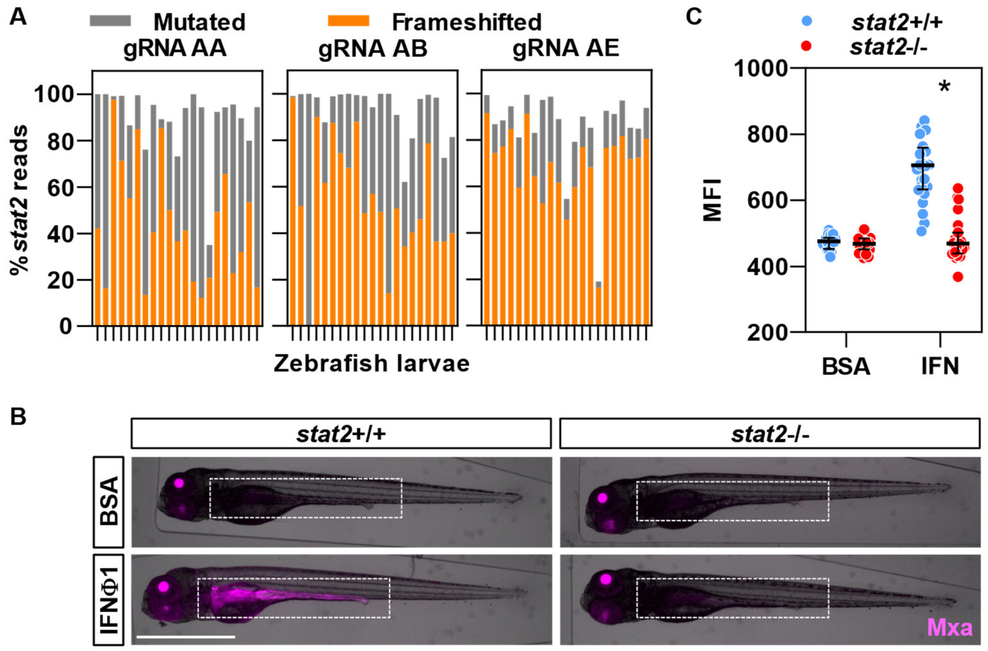


**Figure S8. *stat2* mutagenesis blocks type I interferon signalling in zebrafish, related to Figure 3. (A)** Percentage of mutated (gray) and frameshifted (orange) DNA sequencing reads for each guide RNA (gRNA) target site, predicted using the ampliCan R package, for individual 4-5 day post-fertilization (dpf) zebrafish larvae subjected to yolk sac injection at the one cell stage with three gRNA/Cas9 ribonucleoproteins (RNPs) targeting distinct *stat2* exons. Frameshifting mutations are those in which the length of inserted or deleted nucleotide sequences is not a multiple of three, leading to disruption of the normal reading frame. **(B)** Overlays of brightfield (gray) and fluorescence (magenta) images of *stat2* CRISPant and scrambled RNP injected *Tg(mxa:mCherry)* four dpf larvae 24 hours following intra-coelomic injection of 1 nl of bovine serum albumin (BSA) control or recombinant interferon phi 1 (IFNφ1) protein (1.25 mg/ml). Scale bar = 1 mm. The red signal in the lens is due to the secondary cryaa:DsRed reporter to identify transgene carriers. **(C)** Quantitation of mean fluorescence intensity (MFI) representing Mxa protein expression in the gastrointestinal tract (dashed white outline in **B**). Data points represent individual zebrafish larvae and lines and error bars the median and interquartile range. p values were derived from two-tailed Mann-Whitney tests. * = $p<0.05$. Data are representative of two independent experiments.

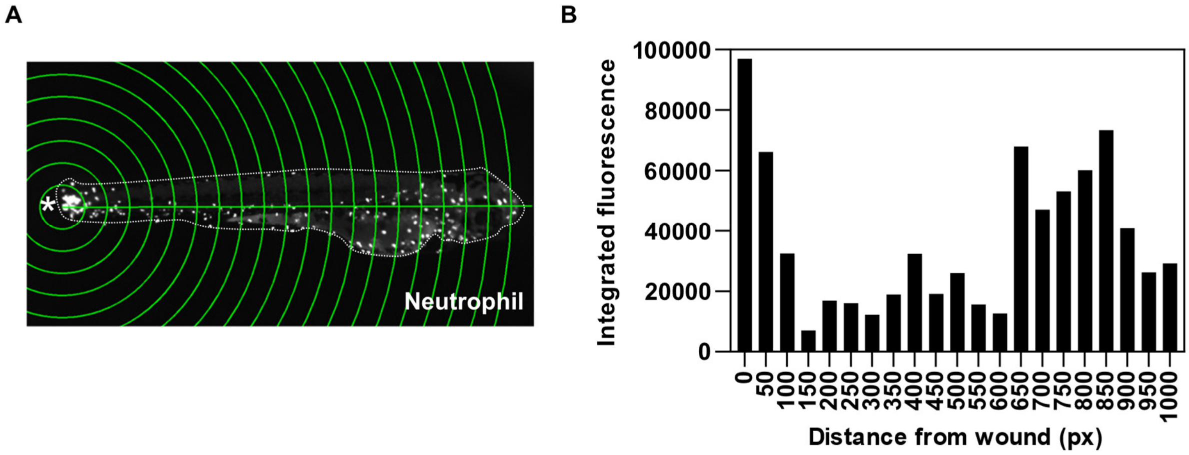


**Figure S9. Quantitation of cellular recruitment to the site of sterile tailfin transection, related to Figures 5 and 6. (A)** Sholl circles (green) superimposed on an image of a three day post-fertilisation *Tg(mpx:eGFP)* zebrafish larva (outlined by the dashed white line) six hours after tailfin transection. The asterisk indicates the tail wound. **(B)** Integrated fluorescence within each circle provides a surrogate for the number of cells at each locus. px = Pixels.

### SUPPLEMENTARY TABLES AND TABLE LEGENDS

**Table S1. Interferon response modules.** Gene composition of the interferon response modules.

**Table S2. Zebrafish lines.** Zebrafish lines used in this study are listed.

| Zebrafish Line | Experiments |
|---|---|
| AB/TL (Tüpfel long fin) referred to as wild type | CRISPant generation<br>Intravenous *M. marinum* infection |
| *Tg(cryaa:DsRed/MXA:mCherry-F)*$^{ump7}$ [22] referred to as *Tg(mxa:mCherry)* | CRISPant generation<br>Confirmation that *stat2* mutagenesis blocks type I interferon signalling |
| *Tg(mpeg1:Gal4-FF)gl25;Tg(UAS-E1b:nfsB.mCherry)c264* [28] referred to as *Tg(mpeg1:mCherry)* | CRISPant generation<br>Quantitation of steady state macrophage numbers<br>Macrophage recruitment to tailfin transection and hindbrain ventricle *M. marinum* infection |
| *Tg(mpx:eGFP)* [29] | CRISPant generation<br>Quantitation of steady state neutrophil numbers<br>Neutrophil recruitment to tailfin transection and hindbrain ventricle *M. marinum* infection |

**Table S3. *stat2* crRNAs, primers and amplicon sequences.** Sequences of the *stat2* crRNAs, Miseq primers and amplicon sequences.

**Table S4. ImageJ macro description.** The workflow and ImageJ functions used to semi-automate generation of single maximum intensity projection montages from the four contiguous z-stacks captured per zebrafish larva using a high-content wide-field fluorescence microscope (Hermes, IDEA Bio-Medical). The macro opens individual images, stitches them together, converts them into a stack, creates a single two-dimensional image with the maximum intensity values and saves the result as a tagged image format (TIF) file in the chosen folder. The saved files can be used for further analysis. The macro is designed to support batch processing, iterating through the specified steps for multiple positions, and concluding with the closure of windows for a clean and efficient workflow.

| Function | Description | Steps |
|---|---|---|
| getDirectory | Prompt the user to select a directory containing images to be processed | Open a dialog box allowing the user to choose a directory |
| getString | Prompt the user to input a keyword representing the specific well position for processing eg A1 | Open an input dialog prompting the user to enter the keyword |
| open | Open an image file for processing | Open individual image files corresponding to the specified keyword and naming convention |
| selectWindow | Select a specific image window for processing | Select the opened image windows according to the specific naming convention for each image in the series |
| run | Execute specific ImageJ commands for image processing | Execute the *"Stack to Images"* command to convert individual z planes into a stack<br>Use the *"Images to Stack"* command with the *"use keep"* option to combine the individual images into a stack while preserving their original order<br>Use the *"Z Project…"* command with the *"projection = [Max Intensity]"* option to create a max intensity projection of the stack |

| | | |
|---|---|---|
| saveAs | Save the processed image with a standardised filename and format | Save the processed image as a TIFF file with a filename constructed using the specified keyword and suffix to indicate processing steps |
| close | Close the active image window or stack after processing | Close the active image windows or stacks to maintain a tidy workspace |

**Table S5. Python script workflow and programming.** The workflow and package functions used to generate measures of total cell numbers and cellular recruitment. BF = Brightfield.

| Workflow | Steps | Package functions used |
|---|---|---|
| get_user_input | Initialize window for user to select BF and fluorescence images | OpenCV: cv2.namedWindow, NumPy: np.array |
| | Open BF image and ask for user-defined start and end point | OpenCV: cv2.imread, cv.2setMouseCallback, cv2.imshow, cv2.waitKey |
| | Return coordinates of selected points | |
| draw_sholl_circles | Calculate distance between selected points | NumPy: np.linalg.norm, np.array |
| | Set distance between Sholl circles | OpenCV: cv2.line, cv2.circle |
| calculate_total_fluorescence | Image preprocessing | OpenCV: cv2.cvtColor, cv2.GaussianBlur, cv2.adaptiveThreshold, SciPy: measure.regionprops, ndimage.label |
| | Calculate integrated fluorescence of entire image | NumPy: np.sum, SciPy: measure.regionprops |
| calculate_fluorescence_per_circle | Mask image based on each Sholl circle | OpenCV: cv2.circle, cv2.bitwise |
| | Compute integrated fluorescence for each Sholl circle | SciPy: measure.regionprops |
| create_csv_files | Organise and save data to CSV files | Pandas: pd.read_csv, pd.DataFrame.to_csv, os: os.path.join, OpenCV: cv2.writer |
| process_image | Coordinate image processing tasks | OpenCV: cv2.imread, cv2.destroyAllWindows |
| | Read input image | OpenCV: cv2.imread |
